# Mental health contact gaps among adults in Peru: a cross-sectional analysis of a nationally representative survey

**DOI:** 10.64898/2026.08.19.26360808

**Authors:** Paulo Ruiz-Grosso, Luis Enrique Macedo Orrego, María Teresa Rivera Encinas, María Soledad Carazas Vera, Diego Mauricio Rodríguez Vargas, Alejandra Arosemena Aliaga, Abel Ampelio Sagástegui Soto, Sonia Zevallos Bustamante

## Abstract

**Objective:** To estimate lifetime and 12-month mental health contact gaps among Peruvian adults with survey-defined mental disorders, and to describe inequalities in contact, perceived need for care, and mental health service use.

**Methods:** We analyzed the information for adults of the 2022 Peruvian National Mental Health Survey, a cross-sectional household survey. The primary outcome was the survey-weighted proportion of adults with a 12-month mental disorder who reported no contact with an included source of mental health-related care during that period; the lifetime contact gap was descriptive. Perceived need was assessed using two derived 12-month perceived-need measures based on direct ENSM variables and service-contact routing items. Analyses incorporated weights, strata, and clusters. Adjusted prevalence ratios were estimated using survey-weighted Poisson models.

**Results:** The dataset contained information on 13,840 individuals; 13,833 had complete survey-design information. Contact-gap denominators were 3,927 for lifetime disorders and 1,649 for 12-month disorders. The lifetime and 12-month contact gaps were 61.0% (95% CI 58.4–63.7) and 84.5% (95% CI 81.5–87.6), respectively. Rural estimates exceeded urban estimates in both periods; after adjustment, poverty and rural residence were associated with the lifetime gap, and extreme poverty with the 12-month gap. Among individuals meeting survey-based criteria for one or more 12-month mental disorders, 37.3% (95% CI 33.4–41.4) reported self-perceived need, whereas 25.6% (95% CI 21.8–29.8) reported that need had been identified by others. Annual psychological and psychiatric service use was 3.7% and 1.1%, respectively.

**Conclusions:** Mental health contact gaps were high, particularly for one or more 12-month mental disorders, and were associated with social and territorial variables. These contact measures do not establish adequate, continuous, or effective treatment; which need to be addressed to understand the impact of the Peruvian mental health reform.

## 1. INTRODUCTION

Mental and substance use disorders are important contributors to the burden of disease in the Americas, accounting for approximately 10% of disability-adjusted life years and 22% of years lived with disability, with broadly similar estimates reported for Latin America (1). This burden is predominantly driven by the prolonged disability associated with mental disorders, which accounted for an estimated 20.6 million disability-adjusted life years in the region in 2019, including 6.9 million attributable to depressive disorders (2). However, drug use disorders have followed a less favorable trajectory, with an estimated 17.7 million people living with these disorders in 2021 and disability-adjusted life-year rates increasing by approximately 5% annually between 2000 and 2021, particularly among young adult men (3). Despite this substantial burden, the health-system response remains limited, as government expenditure on mental health has remained disproportionately low and approximately three quarters of people with moderate-to-severe mental disorders in Latin America do not receive care (1,4).

The treatment gap has been commonly defined as the difference between the prevalence of a disorder and the proportion of affected individuals who receive treatment, or as the proportion of individuals requiring care who do not access it (5). However, because treatment may be defined as anything from any contact with health services to minimally adequate or guideline-concordant effective care, estimates based solely on service contact may substantially underestimate unmet need by overlooking treatment quality, continuity, and adequacy (1,5–7).

International evidence has consistently shown that treatment gaps are larger in low and middle-income countries than in high-income countries (7,8). Recent World Mental Health Survey analyses conceptualize care as a cascade from perceived need and treatment contact through minimally adequate treatment to effective treatment coverage (9). Across 21 countries, only 6.9% of 12-month mental and substance use person-disorders received guideline-consistent effective treatment (9), and fewer than half of cases receiving minimally adequate treatment met criteria for effective treatment coverage (10). Treatment gaps also differ across diagnostic groups, tending to be lower for psychotic disorders than for depressive, anxiety, alcohol use, and other substance use disorders (1,5). More broadly, treatment receipt tends to increase with disorder severity, although substantial unmet need persists even among serious cases (6).

Latin America has historically shown high treatment gaps for mental disorders in a context characterized by fragmented health systems, socioeconomic inequalities, an uneven distribution of specialized services, and persistent demand-side barriers to care (1,11,12). Regional analyses have estimated that approximately three quarters of individuals with moderate-to-severe mental disorders in Latin America do not receive treatment (1). In Peru, previous evidence from the World Mental Health Survey documented substantial unmet need, with treatment receipt increasing with disorder severity but remaining limited even among severe cases (13). These findings provided an essential baseline but were derived from data collected in 2005, before the large-scale expansion of community-based mental health services in Peru and the COVID-19 pandemic, both of which may have subsequently influenced patterns of need, access, and service utilization (13–16). A later population-based study in Metropolitan Lima also documented low annual mental health service access and found that recognition of need did not necessarily translate into service contact (17)

Over the last decade, Peru has expanded community-based mental health services as part of a reform that shifted care toward community and primary-care settings (14,18). Greater service availability, however, does not ensure equitable contact. Recent evidence from Andean Peru suggests that improved geographic proximity to specialized community care can coexist with weak referral pathways, workforce instability, limitations in follow-up, stigma, and tensions between cultural and biomedical understandings of mental health (19). Contemporary population estimates are therefore needed to describe who does and does not report contact with care after this expansion; a cross-sectional survey cannot estimate the effect of the reform itself.

Understanding contact gaps also requires attention to perceived need, which lies between experiencing symptoms and seeking professional help (20). Low perceived need is a frequently reported reason for not initiating care and may reflect mental health literacy, stigma, competing priorities, explanatory models of distress, or the perceived severity of symptoms (12,21). Even when need is recognized, structural and attitudinal barriers may prevent contact or continuity; population analyses should therefore consider perceived need together with the social and geographic distribution of service use (7,9,12,21).

Using the component of the 2022 Peruvian National Mental Health Survey that included the adult population, the primary objective of this study was to estimate the 12-month mental health contact gap among adults meeting survey-based criteria for a mental disorder. Secondary objectives were to describe the lifetime contact gap, differences by diagnostic group, sociodemographic and geographic patterning, perceived need for care, and mental health service use.

## 2. METHODS

### 2.1 Study Design

We conducted a secondary analysis of the adult component of the 2022 Peruvian National Mental Health Survey (Encuesta Nacional de Salud Mental 2022, ENSM 2022). The ENSM 2022 was an observational, cross-sectional, population-based household survey of mental disorders, psychosocial problems, perceived need, and mental health service use. The present analysis focused on mental health contact gaps among adults aged 18–59 years and followed STROBE recommendations for observational studies.

### 2.2 Setting

The ENSM 2022 was conducted in Peru and covered the 25 political regions of the country, including urban and rural areas. The survey was implemented by the Instituto Nacional de Salud Mental “Honorio Delgado – Hideyo Noguchi” and collected data through in-person household interviews during November and December 2022, in the third year after the start of the COVID-19 pandemic. The ENSM 2022 was a national, probabilistic, multistage survey of residents aged two years and older, with diagnostic interviews administered by previously trained field staff.

### 2.3 Participants

The population was adults aged 18–59 years living in non-institutionalized households in Peru. The secondary dataset contained 13,840 records. The survey used a stratified multistage probabilistic household design: census clusters were selected first, dwellings were selected within clusters, and one eligible adult was randomly selected within each household using a Kish table when more than one was eligible.

Participants were eligible if they were adults aged 18–59 years, lived in a non-institutionalized urban or rural residence in Peru, and were available during the data collection period. The survey excluded individuals with hearing impairment, cognitive impairment, neurological conditions, or other communication difficulties that prevented completion of the interview. These exclusion criteria are consistent with the ENSM protocol and final report. The unit of analysis was the individual adult respondent.

### 2.4 Variables

The primary outcome was the 12-month mental health contact gap, defined as the weighted proportion of adults meeting survey-based criteria for a mental disorder in the past 12 months who did not report contact with any included source of mental health-related care during the same period. The annual contact indicator broadly included psychiatrists, psychologists, general physicians or other medical specialists, social workers, counsellors, other mental health professionals, other health professionals, religious or spiritual advisers, and traditional healers. It measures reported contact and not disorder-specific, adequate, continuous, or effective treatment. The lifetime contact gap was a secondary descriptive outcome based on the global survey indicator because provider-level variables did not fully reproduce that indicator.

Mental disorders were assessed using diagnostic variables derived from the adult ENSM 2022 algorithms. Indicators covered any psychiatric disorder and diagnostic groups including anxiety, affective, psychotic, eating, alcohol-use, and other substance-use disorders across lifetime, 12-month, and 30-day reference periods. The supporting variable dictionary documents the variables and coding used in each analysis (Supplementary Table S4).

Mental health service use was assessed by provider and reference period. The present analysis described annual use of psychiatric and psychological services, lifetime use of a telephone-based mental health line, and satisfaction and perceived helpfulness among respondents who used the corresponding professional service.

Sociodemographic and geographic variables were selected a priori and included age, sex, educational level, marital status, poverty based on unmet basic needs, urban or rural residence, natural region, macroregion, and department. Age was analysed continuously in regression models and categorically for descriptive analyses.

Two exploratory indicators of clinical complexity were examined: whether respondents with a 12-month mental disorder also met diagnostic criteria within the past 30 days, and whether they had one versus two or more positive diagnostic domains among five available annual domains: anxiety disorders, affective disorders, psychosis screening, eating disorders, and harmful use or dependence on substances. The 30-day indicator was interpreted as recent diagnostic activity and not as persistence, chronicity, or episode duration. The domain count was considered a partial indicator of diagnostic complexity and not a complete count of mental disorders. Respondents with no positive domain among the five selected domains or incomplete information for any selected domain were not included in the binary domain-count comparison.

### 2.5 Data Sources / Measurement

Data were collected through face-to-face, computer-assisted household interviews using standardized instruments in REDCap. The adult component used a culturally adapted World Health Organization Composite International Diagnostic Interview to generate diagnostic variables based on ICD-10 and DSM-5 criteria.

Two derived 12-month perceived-need measures for mental health care were constructed using direct ENSM perceived-need variables and the SR111 and SR113 routing items. For respondents without annual mental health-service contact, valid responses to the corresponding direct variables were retained. Valid direct responses were also retained when annual contact status was missing. Among respondents reporting annual contact, self-perceived need was coded as present for SR111 responses 1 or 3 and absent for response 2. Need identified by others was coded as present for SR111 responses 2 or 3. When SR111 was 1, SR113 was used to determine whether need had also been identified by someone else: SR113 response 1 was coded as present and response 5 as absent. “Do not know” and non-response categories in the item required by the routing were treated as missing. The corresponding database and analysis variable names are provided in the supporting variable dictionary (Supplementary Table S4). These analytically derived measures were analyzed in the general adult population and among adults meeting survey-based criteria for one or more 12-month mental disorders; their construction should not be interpreted as an official ENSM equivalence between the component items.

Sociodemographic variables were collected through the demographic modules of the survey. Poverty was measured using an unmet-basic-needs classification, and geographic variables were derived from the survey sampling and administrative classification of respondents’ place of residence. Analyses used raked_weight_trimmed as the survey weight.

### 2.6 Bias

Several sources of bias were considered. First, selection bias may have arisen from household non-response or from the exclusion of individuals with hearing impairment, cognitive impairment, neurological conditions, or communication difficulties. The survey attempted to reduce selection bias through probabilistic sampling, household listing procedures, Kish selection within households, and incorporation of the specified survey weight.

Second, information bias may have occurred because both service use and perceived need were self-reported. Recall error may have affected lifetime and 12-month reports of treatment use, and social desirability or non-disclosure may have affected reporting of mental health symptoms, substance use, and service use. The survey attempted to minimize measurement error through standardized interviewer training, computer-assisted data collection, field supervision, and use of validated or culturally adapted instruments.

Third, diagnostic misclassification is possible because psychiatric diagnoses were generated from structured interview algorithms rather than clinical evaluation. However, the use of the WHO-CIDI and diagnostic algorithms based on ICD-10 and DSM-5 criteria improves comparability with other psychiatric epidemiology surveys. The final report also notes that most CIDI modules had acceptable test–retest reliability, although some modules showed lower reliability.

Finally, because the analysis is cross-sectional, the temporality between sociodemographic characteristics, mental disorders, perceived need, and service use cannot be established. Therefore, all associations should be interpreted as prevalence associations rather than causal effects. The survey protocol explicitly identifies the cross-sectional design as a limitation for inferring temporality.

### 2.7 Study Size

The secondary dataset contained 13,840 adult records, of which 13,833 had complete information for the survey design.

For the present secondary analysis, the effective analytic sample was defined by the availability of valid data on diagnostic status, service use, survey weights, and the covariates required for each model. No additional sample size calculation was performed because the analysis used an existing nationally representative survey dataset.

### 2.8 Quantitative Variables

Age was analyzed both as a continuous variable and, for descriptive and subgroup analyses, as a categorical variable using the ENSM adult age groups: 18–29, 30–44, and 45–59 years. Education was categorized as primary education or less, secondary education, and higher education. Marital status was grouped as married/cohabiting, widowed/separated/divorced, and single. Poverty was categorized as non-poor, poor, and extremely poor using the unmet-basic-needs classification. Area of residence was categorized as urban or rural. Natural region was categorized as Coast, Highlands, Jungle, and Metropolitan Lima. Macroregion was categorized as North, Centre, South, East, and Metropolitan Lima, consistent with the survey dictionary.

Diagnostic and contact variables were analysed as binary indicators. Within each diagnostic domain, a contact gap value of 1 indicated no reported contact during the corresponding reference period and a value of 0 indicated reported contact.

### 2.9 Statistical Analysis

Analyses were conducted in Stata 19.5 using Taylor-series linearization. The survey design specified ccluster as the first-stage primary sampling unit, raked_weight_trimmed as the probability weight, and departamentos_si_callao as the stratum; singleton strata were centred. Dwelling and Num_adults were included as nested design variables. Global survey estimates used 13,833 records with complete design information, 25 strata, 2,652 first-stage clusters, and 2,627 design degrees of freedom.

We first described the adult population using weighted proportions and 95% confidence intervals. We then estimated lifetime and 12-month disorder prevalence, service use, and contact gaps overall and by diagnostic group. The 12-month broad-contact definition was primary; alternative provider definitions were examined as sensitivity analyses.

Contact gaps were estimated across sex, age, education, poverty, residence, natural region, macroregion, and department. Group differences were evaluated using design-adjusted Wald tests. Estimates with a coefficient of variation of 30% or greater, fewer than 30 unweighted observations, or fewer than five events or non-events were flagged for cautious interpretation.

Prevalence ratios and 95% confidence intervals were estimated using survey-weighted generalized linear models with a Poisson family and log link. Adjusted models simultaneously included age, sex, education, marital status, poverty, urban or rural residence, and natural region; macroregion was evaluated only in crude sensitivity models. Estimates were interpreted as model-conditional prevalence associations rather than causal effects.

Missing data were handled using complete-case analysis for each outcome-specific model. The number of observations included in each analysis was reported to make denominators transparent. For the derived 12-month perceived-need measures, the primary analysis retained valid direct responses when annual contact status was missing; a sensitivity analysis excluded all observations with missing annual contact status. For the problematic illegal-substance and problematic substance use or dependence indicators, whose unweighted denominators differed between reference periods, a further sensitivity analysis restricted estimation to observations evaluable in both periods. No imputation was performed.

In exploratory analyses restricted to respondents with a 12-month mental disorder, survey-weighted Poisson models with a log link were used to estimate crude prevalence ratios for the annual contact gap according to recent diagnostic activity and according to one versus two or more available diagnostic domains. A further crude model estimated the association between multiple available domains and recent diagnostic activity. These models were not adjusted for sociodemographic covariates and were interpreted as descriptive prevalence associations.

#### Use of generative artificial intelligence

OpenAI ChatGPT and Codex (OpenAI, San Francisco, California; used from August 4 to 19, 2026) were used to assist with drafting and reviewing Stata code and to support structured consistency checks across statistical outputs, tables, and manuscript text. All statistical analyses were run by the investigators in Stata using the ENSM 2022 data. Generative artificial intelligence was not used to generate, impute, or modify participant-level data or to make final analytical or interpretive decisions. The investigators reran the analyses and verified the numerical results against the Stata output.

## 3. RESULTS

### 3.1 Participants

The secondary dataset contained 13,840 adults; 13,833 had complete survey-design information and contributed to global survey estimates. Contact-gap analyses included 3,927 adults who met survey-based criteria for a lifetime disorder and had evaluable lifetime contact, and 1,649 adults who met survey-based criteria for one or more 12-month mental disorders and had evaluable annual contact.

### 3.2 Descriptive Data

Women represented 51.5% of the weighted adult population and the mean age was approximately 36 years. Most participants lived in urban areas (84.4%), and 64.7% were classified as not poor. The prevalence of any psychiatric disorder was 30.9% over the lifetime and 13.4% during the previous 12 months (Table 1).

**Table 1.** Sociodemographic characteristics and prevalence of mental disorders in the ENSM Peru 2022 adult sample, overall and in analytic subpopulations for contact-gap analyses.

| Variable | Full sample |  |  | Lifetime contact-gap analytic subpopulation |  |  | Twelve-month contact-gap analytic subpopulation |  |  |
| --- | --- | --- | --- | --- | --- | --- | --- | --- | --- |
|  | n | Estimate | 95% CI | n | Estimate | 95% CI | n | Estimate | 95% CI |
| Age* | 13833 | 36.05 | 35.7–36.4 | 3927 | 35.85 | 35.25–36.45 | 1649 | 34.52 | 33.49–35.55 |
| Educational attainment | 13815 |  |  | 3919 |  |  | 1645 |  |  |
| No education |  | 3.0 | 2.5–3.6 |  | 2.7 | 1.90–3.83 |  | 2.1 | 1.43–3.20 |
| Preschool |  | 0.20 | 0.1–0.4 |  | 0.14 | 0.04–0.46 |  | 0.05 | 0.01–0.36 |
| Incomplete primary education |  | 6.8 | 6.2–7.4 |  | 7.5 | 6.34–8.98 |  | 7.4 | 5.88–9.20 |
| Complete primary education |  | 9.7 | 9.0–10.5 |  | 8.4 | 7.31–9.71 |  | 7.5 | 5.95–9.43 |
| Incomplete secondary education |  | 8.1 | 7.5–8.8 |  | 8.4 | 7.40–9.56 |  | 9.4 | 7.78–11.28 |
| Complete secondary education |  | 32.7 | 31.3–34.0 |  | 32.7 | 30.41–35.13 |  | 31.0 | 27.09–35.13 |
| Incomplete non-university higher education |  | 5.4 | 4.8–6.0 |  | 5.5 | 4.36–6.81 |  | 7.1 | 5.20–9.60 |
| Complete non-university higher education |  | 11.5 | 10.6–12.5 |  | 10.7 | 9.23–12.39 |  | 9.8 | 7.79–12.29 |
| Incomplete university education |  | 9.3 | 8.4–10.3 |  | 11.0 | 9.26–13.09 |  | 12.9 | 10.20–16.20 |
| Complete university education |  | 11.5 | 10.6–12.5 |  | 10.7 | 9.24–12.36 |  | 9.3 | 7.45–11.66 |
| Postgraduate education |  | 1.7 | 1.1–2.6 |  | 2.0 | 1.01–3.84 |  | 3.3 | 1.40–7.60 |
| Bachelor's degree |  | 0.10 | 0.1–0.3 |  | 0.16 | 0.04–0.59 |  | 0.12 | 0.02–0.67 |
| Marital status | 13627 |  |  | 3903 |  |  | 1636 |  |  |
| Married |  | 24.9 | 23.7–26.2 |  | 22.9 | 20.59–25.41 |  | 21.1 | 17.65–25.02 |
| Separated |  | 7.5 | 6.9–8.3 |  | 8.8 | 7.43–10.40 |  | 8.1 | 6.54–9.99 |
| Divorced |  | 0.96 | 0.7–1.2 |  | 1.0 | 0.65–1.67 |  | 0.81 | 0.29–2.27 |
| Widowed |  | 1.4 | 1.2–1.6 |  | 2.3 | 1.75–2.89 |  | 3.0 | 2.09–4.25 |
| Single |  | 33.3 | 31.9–34.7 |  | 34.6 | 32.24–37.12 |  | 40.4 | 36.33–44.65 |
| Cohabiting |  | 31.9 | 30.6–33.2 |  | 30.4 | 28.04–32.77 |  | 26.6 | 23.27–30.17 |
| Sex | 13692 |  |  | 3920 |  |  | 1645 |  |  |
| Male |  | 48.5 | 47.1–50.0 |  | 47.5 | 44.88–50.08 |  | 43.9 | 39.74–48.19 |
| Female |  | 51.5 | 50.0–52.9 |  | 52.5 | 49.92–55.12 |  | 56.1 | 51.81–60.26 |
| Poverty status | 13833 |  |  | 3927 |  |  | 1649 |  |  |
| Not poor |  | 64.7 | 63.1–66.2 |  | 67.3 | 64.71–69.70 |  | 67.3 | 63.75–70.67 |
| Poor |  | 26.3 | 25.0–27.6 |  | 23.1 | 21.24–25.15 |  | 23.7 | 20.93–26.75 |
| Extreme poverty |  | 9.0 | 8.2–9.9 |  | 9.6 | 8.30–11.09 |  | 9.0 | 7.27–11.04 |
| Area of residence | 13819 |  |  | 3920 |  |  | 1646 |  |  |
|  | n | Estimate | 95% CI | n | Estimate | 95% CI | n | Estimate | 95% CI |
| Urban |  | 84.4 | 82.9–85.8 |  | 88.0 | 86.22–89.66 |  | 89.5 | 87.30–91.37 |
| Rural |  | 15.6 | 14.2–17.1 |  | 12.0 | 10.34–13.78 |  | 10.5 | 8.63–12.70 |
| Natural region | 13815 |  |  | 3920 |  |  | 1645 |  |  |
| Coast |  | 27.4 | 26.0–28.8 |  | 30.8 | 28.32–33.41 |  | 30.8 | 27.41–34.48 |
| Highlands |  | 27.5 | 26.1–28.9 |  | 26.9 | 24.65–29.37 |  | 27.2 | 23.86–30.76 |
| Amazonian region |  | 12.7 | 11.8–13.6 |  | 10.4 | 9.07–11.86 |  | 9.5 | 7.84–11.47 |
| Metropolitan Lima |  | 32.4 | 30.9–34.0 |  | 31.9 | 28.72–35.19 |  | 32.5 | 27.96–37.38 |
| Macroregion | 13823 |  |  | 3922 |  |  | 1646 |  |  |
| North |  | 20.9 | 20.1–21.8 |  | 21.8 | 19.76–23.99 |  | 21.9 | 18.76–25.43 |
| Central |  | 21.9 | 20.6–23.1 |  | 23.0 | 20.93–25.29 |  | 21.7 | 19.01–24.66 |
| South |  | 16.7 | 15.9–17.5 |  | 16.5 | 14.96–18.09 |  | 16.9 | 14.78–19.33 |
| Oriente |  | 8.1 | 7.6–8.6 |  | 6.8 | 5.88–7.95 |  | 7.0 | 5.52–8.76 |
| Metropolitan Lima |  | 32.4 | 30.9–34.0 |  | 31.9 | 28.71–35.18 |  | 32.5 | 27.96–37.37 |
| **Prevalence of mental disorders** |  |  |  |  |  |  |  |  |  |
| **Any mental disorder** |  |  |  |  |  |  |  |  |  |
| Lifetime | 13833 | 30.9 | 29.4–32.4 | 3927 | 100.0 | [—] | 1649 | NR | NR |
| 12 months | 13833 | 13.4 | 12.4–14.4 | 3927 | 43.2 | 40.66–45.66 | 1649 | 100.0 | [—] |
| 1 month | 13833 | 4.8 | 4.1–5.5 | 3927 | 15.2 | 13.34–17.16 | 1649 | 35.2 | 31.30–39.03 |
| **Anxiety disorders** |  |  |  |  |  |  |  |  |  |
| Lifetime | 13833 | 10.1 | 9.2–11.0 | 3927 | 33.1 | 30.57–35.60 | 1649 | 47.7 | 43.52–51.90 |
| 12 months | 13833 | 5.3 | 4.6–5.9 | 3927 | 17.1 | 15.04–19.19 | 1649 | 39.5 | 35.38–43.56 |
| 1 month | 13833 | 2.0 | 1.6–2.5 | 3927 | 6.7 | 5.25–8.11 | 1649 | 15.4 | 12.26–18.55 |
| **Affective disorders** |  |  |  |  |  |  |  |  |  |
| Lifetime | 13833 | 12.8 | 11.8–13.8 | 3927 | 39.7 | 37.13–42.27 | 1649 | 51.4 | 47.26–55.58 |
| 12 months | 13833 | 5.8 | 5.1–6.6 | 3927 | 18.5 | 16.45–20.46 | 1649 | 42.6 | 38.42–46.69 |
| 1 month | 13833 | 2.1 | 1.6–2.5 | 3927 | 6.3 | 5.23–7.37 | 1649 | 14.5 | 12.14–16.94 |
| **Eating disorders** |  |  |  |  |  |  |  |  |  |
| Lifetime | 12610 | 1.2 | 0.8–1.6 | 3761 | 3.8 | 2.55–5.06 | 1577 | 6.0 | 3.44–8.48 |
| 12 months | 12600 | 0.62 | 0.3–0.9 | 3751 | 2.0 | 0.93–3.02 | 1573 | 4.5 | 2.20–6.90 |
| 1 month | 12599 | 0.42 | 0.1–0.7 | 3751 | 1.3 | 0.35–2.31 | 1573 | 3.1 | 0.85–5.29 |
| **Psychosis** |  |  |  |  |  |  |  |  |  |
| Lifetime | 12564 | 1.7 | 1.2–2.2 | 3736 | 5.4 | 3.92–6.81 | 1562 | 7.5 | 4.61–10.38 |
| 12 months | 12564 | 0.47 | 0.3–0.6 | 3736 | 1.5 | 1.01–2.03 | 1562 | 3.5 | 2.32–4.69 |
| **Problematic alcohol use or dependence** |  |  |  |  |  |  |  |  |  |
| Lifetime | 12954 | 8.2 | 7.4–9.0 | 3872 | 26.0 | 23.67–28.36 | 1626 | 23.0 | 19.73–26.35 |
| 12 months | 12954 | 1.7 | 1.4–2.1 | 3872 | 5.3 | 4.26–6.44 | 1626 | 12.5 | 9.97–14.94 |
| **Problematic illegal-substance use or dependence** |  |  |  |  |  |  |  |  |  |
|  | n | Estimate | 95% CI | n | Estimate | 95% CI | n | Estimate | 95% CI |
| Lifetime | 13005 | 0.96 | 0.7–1.2 | 3887 | 3.0 | 2.13–3.83 | 1631 | 3.8 | 2.25–5.35 |
| 12 months | 13832 | 0.20 | 0.1–0.3 | 3926 | 0.65 | 0.33–0.97 | 1649 | 1.5 | 0.76–2.23 |
| **Problematic substance use or dependence** |  |  |  |  |  |  |  |  |  |
| Lifetime | 13684 | 8.8 | 8.0–9.6 | 3927 | 29.3 | 26.87–31.76 | 1649 | 25.6 | 22.21–28.96 |
| 12 months | 13833 | 1.9 | 1.6–2.3 | 3927 | 6.4 | 5.24–7.57 | 1649 | 14.8 | 12.14–17.38 |

Table 1 presents the sociodemographic and diagnostic distributions for the total population and the lifetime and 12-month analytic subpopulations; differences in analytic denominators reflect outcome eligibility and item-specific missingness.

### 3.3 Mental health contact gaps

#### i. Contact gaps by disorder, region, and area of residence

The contact gap was 61.0% (95% CI 58.4–63.7) among adults with a lifetime disorder and 84.5% (95% CI 81.5–87.6) among those with a 12-month disorder. Rural estimates exceeded urban estimates for both lifetime disorders (73.1% vs 59.5%) and 12-month disorders (92.2% vs 83.6%). For lifetime disorders, the Amazonian region had the highest regional estimate (69.2%); regional estimates for 12-month disorders were more similar and less precise. Alternative contact definitions are reported in Supplementary Table S1 (Table 2).

**Table 2.** Contact gaps by mental disorder, period, natural region, and area of residence, ENSM Peru 2022.

**Panel A. Overall estimates and estimates by natural region**
| Disorder / period | n | Total |  | Coast |  | Highlands |  | Amazonian region |  | Metropolitan Lima |  |
| --- | --- | --- | --- | --- | --- | --- | --- | --- | --- | --- | --- |
|  |  | % | 95% CI | % | 95% CI | % | 95% CI | % | 95% CI | % | 95% CI |
| Any mental disorder |  |  |  |  |  |  |  |  |  |  |  |
| Lifetime | 3927 | 61.0 | 58.4–63.7 | 60.1 | 56.5–63.8 | 62.8 | 58.7–66.9 | 69.2 | 64.0–74.5 | 58.6 | 52.3–64.9 |
| 12 months | 1649 | 84.5 | 81.5–87.6 | 83.8 | 79.9–87.7 | 87.1 | 83.3–90.8 | 83.8 | 77.5–90.1 | 82.9 | 75.0–90.9 |
| Anxiety disorders |  |  |  |  |  |  |  |  |  |  |  |
| Lifetime | 1316 | 49.0 | 44.4–53.7 | 51.1 | 44.7–57.5 | 53.9 | 46.7–61.1 | 58.3 | 49.1–67.4 | 41.5 | 30.8–52.3 |
| 12 months | 666 | 80.9 | 75.4–86.4* | 80.1 | 73.0–87.1* | 85.4 | 79.1–91.7* | 87.2 | 78.7–95.7 | 75.9 | 61.7–90.0* |
| Affective disorders |  |  |  |  |  |  |  |  |  |  |  |
| Lifetime | 1672 | 52.1 | 48.2–55.9 | 56.0 | 50.1–61.9 | 53.4 | 47.2–59.5 | 58.5 | 50.5–66.6 | 44.9 | 36.0–53.8 |
| 12 months | 732 | 80.9 | 76.1–85.7 | 83.3 | 77.5–89.1 | 81.0 | 75.1–86.9 | 77.0 | 66.2–87.8 | 79.5 | 67.1–92.0 |
| Eating disorders |  |  |  |  |  |  |  |  |  |  |  |
| Lifetime | 120 | 57.2 | 42.0–72.5* | 45.6 | 28.8–62.3* | 79.9 | 60.0–99.8* | 37.2 | 11.8–62.7 | 55.2 | 24.2–86.2* |
| 12 months | 54 | 73.6 | 52.6–94.6* | 70.9 | 48.0–93.9* | 90.6 | 73.4–107.8* | 58.5 | 15.6–101.5 | 60.9 | 13.0–108.8* |
| Psychosis |  |  |  |  |  |  |  |  |  |  |  |
| Lifetime | 176 | 58.3 | 45.3–71.3* | 51.7 | 36.3–67.2* | 62.2 | 48.9–75.4* | 50.0 | 21.6–78.5 | 62.2 | 34.7–89.8* |
| 12 months | 56 | 77.1 | 62.6–91.7* | 81.4 | 64.1–98.7* | 94.3 | 85.3–103.2* | 73.0 | 35.0–111.0 | 36.5 | -16.7–89.8* |
| Problematic alcohol use or dependence |  |  |  |  |  |  |  |  |  |  |  |
| Lifetime | 850 | 66.5 | 61.9–71.1* | 65.7 | 58.3–73.1* | 62.1 | 55.2–69.0* | 75.3 | 66.3–84.3 | 68.1 | 56.4–79.8* |
| 12 months | 186 | 84.9 | 75.7–94.1* | 89.9 | 80.4–99.4* | 89.1 | 80.2–98.0* | 88.1 | 76.5–99.7 | 73.2 | 44.7–101.8* |
| Problematic substance use or dependence |  |  |  |  |  |  |  |  |  |  |  |
| Lifetime | 983 | 63.1 | 58.6–67.6 | 62.3 | 55.4–69.3 | 59.6 | 53.0–66.1 | 72.9 | 64.3–81.4 | 63.2 | 52.3–74.1 |
| 12 months | 231 | 82.7 | 74.6–90.8 | 90.4 | 82.6–98.2* | 84.5 | 74.5–94.5* | 84.1 | 71.9–96.4* | 72.3 | 48.1–96.4* |
| Problematic illegal-substance use or dependence |  |  |  |  |  |  |  |  |  |  |  |
| Lifetime | 102 | 32.6 | 17.8–47.4* | 26.9 | 11.2–42.6* | 21.7 | 4.9–38.4* | 16.8 | -10.5–44.1 | 43.1 | 14.3–71.9* |
| 12 months | 28 | 64.8 | 37.5–92.0* | 85.6 | 64.1–107.2* | 63.5 | 6.6–120.5* | 59.6 | 3.2–116.0 | 48.6 | -4.1–101.4* |

**Panel B. Estimates by area of residence**
| Disorder / period | Urban |  | Rural |  |
| --- | --- | --- | --- | --- |
|  | % | 95% CI | % | 95% CI |
| **Any mental disorder** |  |  |  |  |
| Lifetime | 59.5 | 56.6–62.4 | 73.1 | 68.4–77.8 |
| 12 months | 83.6 | 80.2–87.0 | 92.2 | 88.3–96.0 |
| **Anxiety disorders** |  |  |  |  |
| Lifetime | 47.7 | 42.7–52.7 | 62.0 | 52.0–71.9 |
| 12 months | 79.7 | 73.7–85.8 | 89.7 | 81.3–98.1 |
| **Affective disorders** |  |  |  |  |
|  | % | 95% CI | % | 95% CI |
| Lifetime | 49.8 | 45.6–54.0 | 68.7 | 61.8–75.6 |
| 12 months | 79.9 | 74.6–85.2 | 89.1 | 83.1–95.2 |
| **Eating disorders** |  |  |  |  |
| Lifetime | 54.6 | 37.9–71.3 | 100.0 | —† |
| 12 months | 72.7 | 51.0–94.5 | 100.0 | —† |
| **Psychosis** |  |  |  |  |
| Lifetime | 57.5 | 42.7–72.2 | 61.1 | 46.8–75.4 |
| 12 months | 75.5 | 58.8–92.2 | 85.0 | 61.5–108.4 |
| **Problematic alcohol use or dependence** |  |  |  |  |
| Lifetime | 65.4 | 60.3–70.5 | 76.0 | 68.0–84.1 |
| 12 months | 83.3 | 73.0–93.5 | 98.0 | 94.1–101.9 |
| **Problematic substance use or dependence** |  |  |  |  |
| Lifetime | 61.8 | 56.9–66.7 | 74.9 | 66.7–83.1 |
| 12 months | 81.0 | 72.0–89.9 | 98.2 | 94.8–101.7 |
| **Problematic illegal-substance use or dependence** |  |  |  |  |
| Lifetime | 32.1 | 16.9–47.3 | 53.0 | 1.5–104.6 |
| 12 months | 63.6 | 35.6–91.6 | 100.0 | —† |

Values are weighted percentages without contact with any included source among participants meeting survey-based criteria for the disorder during the corresponding period. These estimates describe contact and do not establish that care was adequate, continuous, or effective. The 95% CIs account for the complex survey design. Urban/rural estimates used raked\_weight\_trimmed, departamentos\_si\_callao strata, ccluster primary sampling units, and singleunit(centered). \* Estimate flagged for cautious interpretation because of small effective sample size and/or high imprecision. † The CI was non-informative because no variation was observed (estimate 100%; standard error 0). ENSM, National Mental Health Survey. The 95% CIs in this table were computed on the linear (Wald) scale using Taylor-series linearization and were not bounded by a logit transformation; in sparse strata this can yield limits below 0% or above 100%, which should be read as non-informative bounds rather than as attainable prevalences.

Contact gaps varied across diagnostic groups and were high for alcohol- and other substance-use disorders. Estimates for psychotic and eating disorders and for several rural or regional strata were imprecise because of small effective samples or sparse events and should be interpreted cautiously (Table 2).

#### ii. Sociodemographic factors associated with contact gaps

Crude prevalence ratios are presented in Table 3. Poverty and rural residence showed the most consistent crude associations with higher lifetime and 12-month contact gaps, whereas several education, marital-status, and regional associations attenuated after adjustment.

**Table 3.**
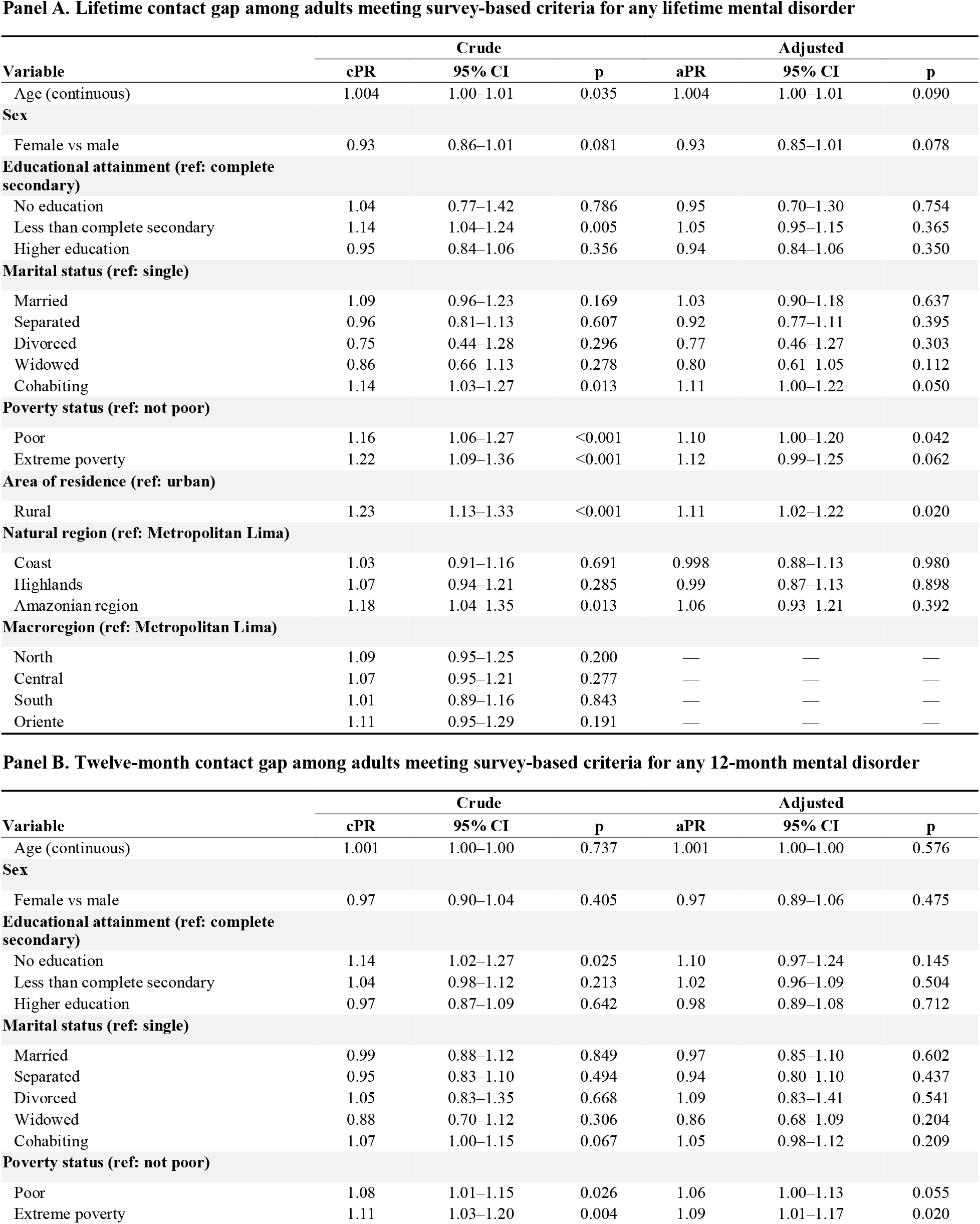
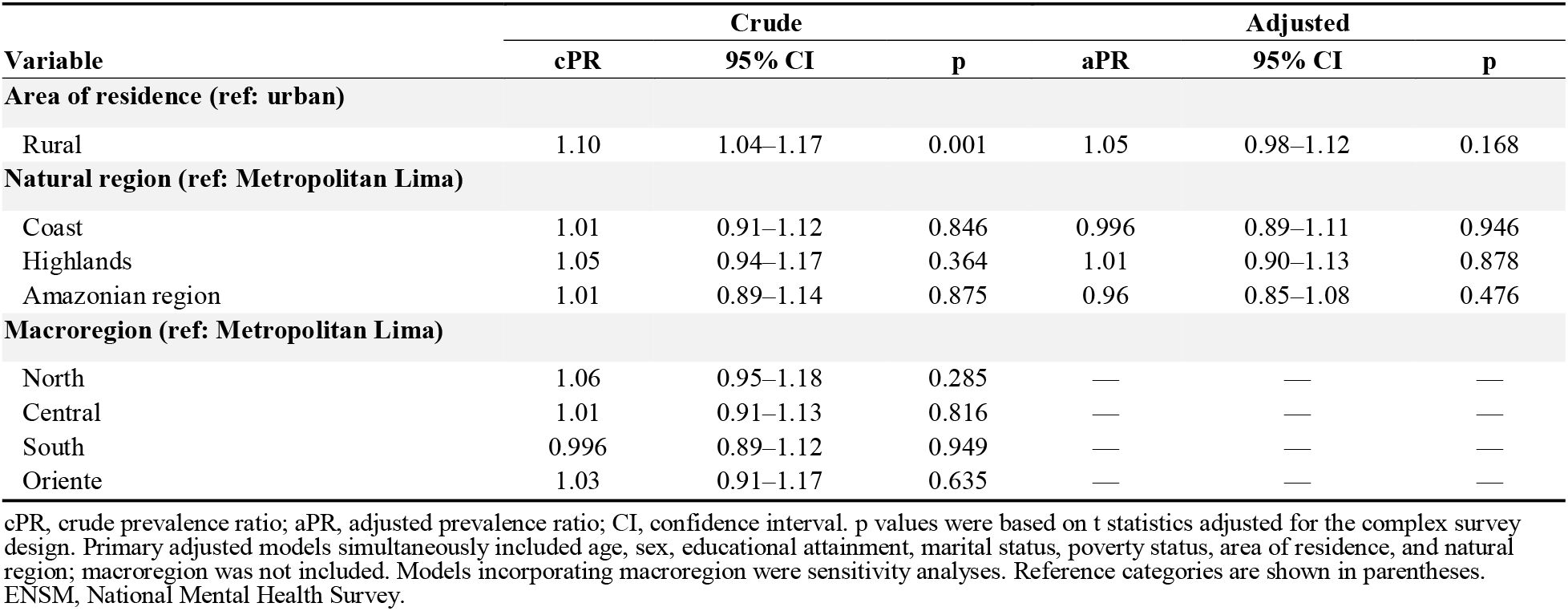
Factors associated with mental health contact gaps over the lifetime and previous 12 months, ENSM Peru 2022.

**Panel A. Lifetime contact gap among adults meeting survey-based criteria for any lifetime mental disorder**
| Variable | Crude |  |  | Adjusted |  |  |
| --- | --- | --- | --- | --- | --- | --- |
|  | cPR | 95% CI | p | aPR | 95% CI | p |
| Age (continuous) | 1.004 | 1.00–1.01 | 0.035 | 1.004 | 1.00–1.01 | 0.090 |
| **Sex** |  |  |  |  |  |  |
| Female vs male | 0.93 | 0.86–1.01 | 0.081 | 0.93 | 0.85–1.01 | 0.078 |
| **Educational attainment (ref: complete secondary)** |  |  |  |  |  |  |
| No education | 1.04 | 0.77–1.42 | 0.786 | 0.95 | 0.70–1.30 | 0.754 |
| Less than complete secondary | 1.14 | 1.04–1.24 | 0.005 | 1.05 | 0.95–1.15 | 0.365 |
| Higher education | 0.95 | 0.84–1.06 | 0.356 | 0.94 | 0.84–1.06 | 0.350 |
| **Marital status (ref: single)** |  |  |  |  |  |  |
| Married | 1.09 | 0.96–1.23 | 0.169 | 1.03 | 0.90–1.18 | 0.637 |
| Separated | 0.96 | 0.81–1.13 | 0.607 | 0.92 | 0.77–1.11 | 0.395 |
| Divorced | 0.75 | 0.44–1.28 | 0.296 | 0.77 | 0.46–1.27 | 0.303 |
| Widowed | 0.86 | 0.66–1.13 | 0.278 | 0.80 | 0.61–1.05 | 0.112 |
| Cohabiting | 1.14 | 1.03–1.27 | 0.013 | 1.11 | 1.00–1.22 | 0.050 |
| **Poverty status (ref: not poor)** |  |  |  |  |  |  |
| Poor | 1.16 | 1.06–1.27 | <0.001 | 1.10 | 1.00–1.20 | 0.042 |
| Extreme poverty | 1.22 | 1.09–1.36 | <0.001 | 1.12 | 0.99–1.25 | 0.062 |
| **Area of residence (ref: urban)** |  |  |  |  |  |  |
| Rural | 1.23 | 1.13–1.33 | <0.001 | 1.11 | 1.02–1.22 | 0.020 |
| **Natural region (ref: Metropolitan Lima)** |  |  |  |  |  |  |
| Coast | 1.03 | 0.91–1.16 | 0.691 | 0.998 | 0.88–1.13 | 0.980 |
| Highlands | 1.07 | 0.94–1.21 | 0.285 | 0.99 | 0.87–1.13 | 0.898 |
| Amazonian region | 1.18 | 1.04–1.35 | 0.013 | 1.06 | 0.93–1.21 | 0.392 |
| **Macroregion (ref: Metropolitan Lima)** |  |  |  |  |  |  |
| North | 1.09 | 0.95–1.25 | 0.200 | — | — | — |
| Central | 1.07 | 0.95–1.21 | 0.277 | — | — | — |
| South | 1.01 | 0.89–1.16 | 0.843 | — | — | — |
| Oriente | 1.11 | 0.95–1.29 | 0.191 | — | — | — |

**Panel B. Twelve-month contact gap among adults meeting survey-based criteria for any 12-month mental disorder**
| Variable | Crude |  |  | Adjusted |  |  |
| --- | --- | --- | --- | --- | --- | --- |
|  | cPR | 95% CI | p | aPR | 95% CI | p |
| Age (continuous) | 1.001 | 1.00–1.00 | 0.737 | 1.001 | 1.00–1.00 | 0.576 |
| **Sex** |  |  |  |  |  |  |
| Female vs male | 0.97 | 0.90–1.04 | 0.405 | 0.97 | 0.89–1.06 | 0.475 |
| **Educational attainment (ref: complete secondary)** |  |  |  |  |  |  |
| No education | 1.14 | 1.02–1.27 | 0.025 | 1.10 | 0.97–1.24 | 0.145 |
| Less than complete secondary | 1.04 | 0.98–1.12 | 0.213 | 1.02 | 0.96–1.09 | 0.504 |
| Higher education | 0.97 | 0.87–1.09 | 0.642 | 0.98 | 0.89–1.08 | 0.712 |
| **Marital status (ref: single)** |  |  |  |  |  |  |
| Married | 0.99 | 0.88–1.12 | 0.849 | 0.97 | 0.85–1.10 | 0.602 |
| Separated | 0.95 | 0.83–1.10 | 0.494 | 0.94 | 0.80–1.10 | 0.437 |
| Divorced | 1.05 | 0.83–1.35 | 0.668 | 1.09 | 0.83–1.41 | 0.541 |
| Widowed | 0.88 | 0.70–1.12 | 0.306 | 0.86 | 0.68–1.09 | 0.204 |
| Cohabiting | 1.07 | 1.00–1.15 | 0.067 | 1.05 | 0.98–1.12 | 0.209 |
| **Poverty status (ref: not poor)** |  |  |  |  |  |  |
| Poor | 1.08 | 1.01–1.15 | 0.026 | 1.06 | 1.00–1.13 | 0.055 |
| Extreme poverty | 1.11 | 1.03–1.20 | 0.004 | 1.09 | 1.01–1.17 | 0.020 |
|  | cPR | 95% CI | p | aPR | 95% CI | p |
| **Area of residence (ref: urban)** |  |  |  |  |  |  |
| Rural | 1.10 | 1.04–1.17 | 0.001 | 1.05 | 0.98–1.12 | 0.168 |
| **Natural region (ref: Metropolitan Lima)** |  |  |  |  |  |  |
| Coast | 1.01 | 0.91–1.12 | 0.846 | 0.996 | 0.89–1.11 | 0.946 |
| Highlands | 1.05 | 0.94–1.17 | 0.364 | 1.01 | 0.90–1.13 | 0.878 |
| Amazonian region | 1.01 | 0.89–1.14 | 0.875 | 0.96 | 0.85–1.08 | 0.476 |
| **Macroregion (ref: Metropolitan Lima)** |  |  |  |  |  |  |
| North | 1.06 | 0.95–1.18 | 0.285 | — | — | — |
| Central | 1.01 | 0.91–1.13 | 0.816 | — | — | — |
| South | 0.996 | 0.89–1.12 | 0.949 | — | — | — |
| Oriente | 1.03 | 0.91–1.17 | 0.635 | — | — | — |

cPR, crude prevalence ratio; aPR, adjusted prevalence ratio; CI, confidence interval. p values were based on t statistics adjusted for the complex survey design. Primary adjusted models simultaneously included age, sex, educational attainment, marital status, poverty status, area of residence, and natural region; macroregion was not included. Models incorporating macroregion were sensitivity analyses. Reference categories are shown in parentheses. ENSM, National Mental Health Survey.

In adjusted models, poverty (PR 1.10, 95% CI 1.00–1.20) and rural residence (PR 1.11, 95% CI 1.02–1.22) were associated with a higher lifetime contact gap. For the 12-month contact gap, extreme poverty remained associated with a higher prevalence (PR 1.09, 95% CI 1.01–1.17); other adjusted sociodemographic and regional estimates included the null (Table 3).

### 3.4 Exploratory indicators of clinical complexity

Among 1,708 adults meeting survey-based criteria for one or more 12-month mental disorders, 35.7% (95% CI 31.8–39.8) also met criteria within the previous 30 days. The annual contact gap was 80.0% among those with recent diagnostic activity and 87.0% among those without it (crude PR 0.92, 95% CI 0.85–1.00). Among 1,367 respondents in the domain-count comparison (1,361 with an evaluable annual contact gap), the contact gap was 71.5% for those with two or more available diagnostic domains and 87.3% for those with one domain (crude PR 0.82, 95% CI 0.72–0.93). These exploratory indicators do not measure severity, persistence, or comprehensive clinical need (Supplementary Table S3).

### 3.5 Perceived need for mental health care

#### i. Perceived need for mental health care by disorder, region, and area of residence

In the general adult population, 14.3% (95% CI 13.3–15.4) reported self-perceived need and 9.0% (95% CI 8.0–10.0) had need identified by others. Among adults meeting survey-based criteria for one or more 12-month mental disorders, the corresponding estimates were 37.3% (95% CI 33.4–41.4) and 25.6% (95% CI 21.8–29.8). Within this survey-defined subgroup, self-perceived need was 39.0% in urban and 23.4% in rural residents, while need identified by others was 27.3% and 12.0%, respectively (Table 4).

**Table 4.** Twelve-month perceived need for mental health care according to a derived measure, ENSM Peru 2022.

**Panel A. Overall estimates and estimates by natural region**
| Type of perceived need | Total |  | Coast |  | Highlands |  | Amazonian region |  | Metropolitan Lima |  |
| --- | --- | --- | --- | --- | --- | --- | --- | --- | --- | --- |
|  | % | 95% CI | % | 95% CI | % | 95% CI | % | 95% CI | % | 95% CI |
| **Perceived need (general adult population)** |  |  |  |  |  |  |  |  |  |  |
| Self-perceived | 14.3 | 13.3–15.4 | 15.8 | 14.2–17.6 | 12.5 | 11.0–14.1 | 9.7 | 7.7–12.1 | 16.6 | 14.1–19.4 |
| Identified by others | 9.0 | 8.0–10.0 | 8.9 | 7.7–10.1 | 7.4 | 6.1–8.9 | 5.5 | 3.8–7.7 | 11.9 | 9.6–14.7 |
| **Perceived need (adults meeting survey-based criteria for ≥1 12-month mental disorder)** |  |  |  |  |  |  |  |  |  |  |
| Self-perceived | 37.3 | 33.4–41.4 | 38.9 | 33.3–44.7 | 33.4 | 27.4–40.1 | 30.9 | 23.1–40.0 | 41.2 | 32.1–50.9 |
| Identified by others | 25.6 | 21.8–29.8 | 25.6 | 20.8–31.2 | 23.8 | 17.0–32.2 | 15.7 | 10.3–23.2 | 30.1 | 21.8–39.9 |
| **Anxiety disorders** |  |  |  |  |  |  |  |  |  |  |
| Self-perceived | 44.4 | 37.7–51.2 | 46.7 | 37.9–55.7 | 35.2 | 26.9–44.7 | 24.7 | 15.2–37.5 | 53.7 | 37.2–69.4 |
| Identified by others | 26.5 | 20.8–33.1 | 26.6 | 19.4–35.4 | 18.4 | 12.2–26.9 | 21.4 | 11.5–36.4 | 33.3 | 20.2–49.6 |
| **Affective disorders** |  |  |  |  |  |  |  |  |  |  |
| Self-perceived | 43.8 | 38.3–49.5 | 41.8 | 32.9–51.3 | 40.8 | 33.7–48.4 | 42.6 | 29.2–57.3 | 49.1 | 36.7–61.6 |
| Identified by others | 29.6 | 24.6–35.2 | 28.4 | 20.9–37.3 | 24.6 | 18.4–32.0 | 16.1 | 9.1–26.9 | 39.3 | 27.3–52.7 |
| **Eating disorders** |  |  |  |  |  |  |  |  |  |  |
| Self-perceived | 67.5 | 42.9–85.2 | 70.7 | 45.3–87.5* | 95.8 | 77.6–99.3* | 41.5 | 8.9–83.6* | 39.5 | 10.5–78.4* |
| Identified by others | 62.0 | 39.1–80.5 | 45.0 | 23.5–68.5* | 75.5 | 30.7–95.6* | 17.6 | 2.1–67.4* | 73.3 | 37.3–92.7* |
| **Psychosis** |  |  |  |  |  |  |  |  |  |  |
| Self-perceived | 25.2 | 13.5–42.1 | 22.4 | 7.5–50.6* | 19.6 | 7.1–43.6* | 29.8 | 6.5–72.4* | 37.2 | 7.2–82.0* |
| Identified by others | 28.2 | 14.8–46.9 | 23.4 | 8.1–51.6* | 31.3 | 12.3–59.7* | 0.0 | NE* | 63.5 | 18.5–93.0* |
| **Problematic alcohol use or dependence** |  |  |  |  |  |  |  |  |  |  |
| Self-perceived | 28.7 | 21.0–37.9 | 28.6 | 15.9–45.9 | 33.9 | 22.2–47.9 | 31.9 | 16.9–51.8* | 21.1 | 7.8–45.8* |
| Identified by others | 23.2 | 14.8–34.4 | 26.9 | 13.8–45.9 | 21.1 | 12.2–33.9 | 9.8 | 2.7–30.3* | 28.2 | 9.1–60.8* |
| **Problematic substance use or dependence** |  |  |  |  |  |  |  |  |  |  |
| Self-perceived | 30.1 | 22.9–38.5 | 26.7 | 15.8–41.4 | 40.3 | 28.3–53.7 | 32.2 | 18.3–50.1 | 20.7 | 8.7–41.5 |
| Identified by others | 25.1 | 17.3–34.8 | 28.7 | 16.6–44.9 | 22.8 | 13.4–36.0 | 11.5 | 3.9–29.6* | 30.2 | 12.3–57.1 |
| **Problematic illegal-substance use or dependence** |  |  |  |  |  |  |  |  |  |  |
| Self-perceived | 43.9 | 22.2–68.2* | 20.1 | 6.1–49.6* | 88.7 | 43.8–98.8* | 29.6 | 3.9–81.4* | 30.5 | 6.7–72.9* |
| Identified by others | 60.0 | 36.5–79.6* | 61.1 | 27.3–86.7* | 47.3 | 7.7–90.6* | 62.7 | 16.5–93.5* | 68.6 | 25.9–93.2* |

**Panel B. Estimates by area of residence**
| Type of perceived need | Urban |  | Rural |  |
| --- | --- | --- | --- | --- |
|  | % | 95% CI | % | 95% CI |
| **Perceived need (general adult population)** |  |  |  |  |
| Self-perceived | 15.7 | 14.5–17.0 | 7.1 | 5.7–8.7 |
| Identified by others | 10.0 | 8.9–11.2 | 3.8 | 2.9–4.9 |
|  | % | 95% CI | % | 95% CI |
| **Perceived need (adults meeting survey-based criteria for ≥1 12-month mental disorder)** |  |  |  |  |
| Self-perceived | 39.0 | 34.7–43.5 | 23.4 | 17.6–30.5 |
| Identified by others | 27.3 | 23.2–32.0 | 12.0 | 7.9–17.6 |
| **Anxiety disorders** |  |  |  |  |
| Self-perceived | 46.6 | 39.3–53.9 | 25.6 | 15.0–40.2 |
| Identified by others | 28.6 | 22.3–35.8 | 11.0 | 5.5–20.9 |
| **Affective disorders** |  |  |  |  |
| Self-perceived | 46.2 | 40.1–52.4 | 23.8 | 15.6–34.6 |
| Identified by others | 31.6 | 26.0–37.7 | 13.5 | 7.9–22.2 |
| **Eating disorders** |  |  |  |  |
| Self-perceived | 69.8 | 44.6–86.9 | 0.0 | NE* |
| Identified by others | 64.0 | 41.1–81.9 | 0.0 | NE* |
| **Psychosis** |  |  |  |  |
| Self-perceived | 24.7 | 12.2–43.7 | 28.4 | 7.4–66.3* |
| Identified by others | 30.8 | 15.8–51.4 | 10.8 | 2.2–39.7* |
| **Problematic alcohol use or dependence** |  |  |  |  |
| Self-perceived | 27.7 | 19.5–37.7 | 36.9 | 18.5–60.1 |
| Identified by others | 24.2 | 14.9–36.7 | 16.9 | 6.1–38.9 |
| **Problematic substance use or dependence** |  |  |  |  |
| Self-perceived | 29.8 | 22.1–38.9 | 33.4 | 16.5–56.0 |
| Identified by others | 26.3 | 17.8–37.0 | 15.3 | 5.5–35.8 |
| **Problematic illegal-substance use or dependence** |  |  |  |  |
| Self-perceived | 48.2 | 24.5–72.8* | 0.0 | NE* |
| Identified by others | 59.5 | 34.7–80.2* | 64.8 | 10.1–96.8* |

Results were weighted for the complex ENSM Peru 2022 survey design. The derived measure retained valid direct responses among participants without annual contact; among participants with annual contact, self-perceived need was based on SR111 and need identified by others on SR111 and, when SR111 = 1, SR113. 'Do not know' and nonresponse were treated as missing. The 95% CIs were bounded using a logit transformation. \* Unweighted n < 30 or fewer than five events or non-events; interpret cautiously. NE, not estimable at a 0/1 boundary. ENSM, National Mental Health Survey.

Perceived-need estimates varied by diagnostic group, but several regional and residence- stratified estimates for eating disorders, psychosis, and illegal-substance use disorders were based on sparse data and were flagged for cautious interpretation. Excluding observations with missing annual contact status changed the four main prevalence estimates by no more than 0.19 percentage points; the large change in one illegal-substance-use rural cell reflected a comparison of two versus one unweighted observation and was not interpreted (Table 4; sensitivity analysis).

#### ii. Sociodemographic variables associated with perceived need for mental health care

In the general population, adjusted self-perceived need was higher among women (PR 1.77, 95% CI 1.51–2.08) and lower among adults in extreme poverty (PR 0.66, 95% CI 0.51–0.84) and rural residents (PR 0.58, 95% CI 0.46–0.74). Need identified by others was higher among women (PR 1.30, 95% CI 1.04–1.63) and lower among adults in extreme poverty (PR 0.66, 95% CI 0.46–0.93), rural residents (PR 0.54, 95% CI 0.39–0.74), and residents of the Amazonian region (PR 0.62, 95% CI 0.40–0.95). Among adults meeting survey-based criteria for one or more 12-month mental disorders, self-perceived need was higher among women (PR 1.53, 95% CI 1.19–1.98) and those with higher education (PR 1.43, 95% CI 1.11–1.85); need identified by others was lower among those in extreme poverty (PR 0.51, 95% CI 0.31–0.83) and higher among widowed than single adults (PR 1.91, 95% CI 1.08–3.36). For the latter outcome, the adjusted rural estimate (PR 0.66, 95% CI 0.42–1.05) and crude Oriente macroregion estimate (PR 0.62, 95% CI 0.37–1.06) included the null in the primary analysis and crossed the conventional significance threshold only when observations with missing contact status were excluded; they were treated as unstable rather than robust associations (Table 5).

**Table 5.** Factors associated with twelve-month perceived need for mental health care according to a derived measure, ENSM Peru 2022.

**Panel A. General adult population: self-perceived need**
| Variable | Crude |  |  | Adjusted |  |  |
| --- | --- | --- | --- | --- | --- | --- |
|  | cPR | 95% CI | p | aPR | 95% CI | p |
| Age (continuous) | 0.99 | 0.98–0.99 | <0.001 | 0.99 | 0.99–1.00 | 0.069 |
| **Sex** |  |  |  |  |  |  |
| Female vs male | 1.69 | 1.44–1.98 | <0.001 | 1.77 | 1.51–2.08 | <0.001 |
| **Educational attainment (ref: complete secondary)** |  |  |  |  |  |  |
| No education | 0.29 | 0.15–0.57 | <0.001 | 0.30 | 0.16–0.57 | <0.001 |
| Less than complete secondary | 0.72 | 0.61–0.86 | <0.001 | 0.89 | 0.74–1.06 | 0.192 |
| Higher education | 1.17 | 0.97–1.40 | 0.094 | 1.13 | 0.94–1.35 | 0.189 |
| **Marital status (ref: single)** |  |  |  |  |  |  |
| Married | 0.64 | 0.53–0.79 | <0.001 | 0.71 | 0.58–0.87 | <0.001 |
| Separated | 0.84 | 0.67–1.04 | 0.114 | 0.84 | 0.68–1.05 | 0.125 |
| Divorced | 1.97 | 1.28–3.01 | 0.002 | 1.76 | 1.15–2.69 | 0.009 |
| Widowed | 1.20 | 0.82–1.74 | 0.347 | 1.26 | 0.85–1.86 | 0.250 |
| Cohabiting | 0.79 | 0.66–0.95 | 0.012 | 0.87 | 0.72–1.04 | 0.116 |
| **Poverty status (ref: not poor)** |  |  |  |  |  |  |
| Poor | 0.79 | 0.68–0.93 | 0.004 | 0.95 | 0.81–1.12 | 0.541 |
| Extreme poverty | 0.47 | 0.37–0.60 | <0.001 | 0.66 | 0.51–0.84 | <0.001 |
| **Area of residence (ref: urban)** |  |  |  |  |  |  |
| Rural | 0.45 | 0.36–0.57 | <0.001 | 0.58 | 0.46–0.74 | <0.001 |
| **Natural region (ref: Metropolitan Lima)** |  |  |  |  |  |  |
| Coast | 0.96 | 0.79–1.16 | 0.643 | 1.04 | 0.86–1.25 | 0.700 |
| Highlands | 0.75 | 0.62–0.92 | 0.006 | 0.98 | 0.80–1.19 | 0.830 |
| Amazonian region | 0.58 | 0.44–0.77 | <0.001 | 0.78 | 0.59–1.03 | 0.083 |
| **Macroregion (ref: Metropolitan Lima)** |  |  |  |  |  |  |
| North | 0.76 | 0.61–0.96 | 0.019 | — | — | — |
| Central | 0.94 | 0.78–1.15 | 0.568 | — | — | — |
| South | 0.76 | 0.62–0.94 | 0.009 | — | — | — |
| Oriente | 0.62 | 0.44–0.87 | 0.006 | — | — | — |

**Panel B. General adult population: need identified by others**
| Variable | Crude |  |  | Adjusted |  |  |
| --- | --- | --- | --- | --- | --- | --- |
|  | cPR | 95% CI | p | aPR | 95% CI | p |
| Age (continuous) | 0.98 | 0.97–0.99 | <0.001 | 0.99 | 0.98–1.00 | 0.110 |
| **Sex** |  |  |  |  |  |  |
| Female vs male | 1.26 | 1.00–1.57 | 0.046 | 1.30 | 1.04–1.63 | 0.022 |
| **Educational attainment (ref: complete secondary)** |  |  |  |  |  |  |
| No education | 0.45 | 0.20–1.01 | 0.052 | 0.64 | 0.27–1.52 | 0.306 |
| Less than complete secondary | 0.69 | 0.54–0.89 | 0.004 | 0.99 | 0.77–1.27 | 0.919 |
| Higher education | 1.12 | 0.87–1.46 | 0.381 | 1.14 | 0.87–1.51 | 0.346 |
| **Marital status (ref: single)** |  |  |  |  |  |  |
| Married | 0.48 | 0.35–0.67 | <0.001 | 0.57 | 0.40–0.81 | 0.002 |
| Separated | 0.81 | 0.58–1.13 | 0.213 | 0.89 | 0.61–1.28 | 0.518 |
| Divorced | 0.93 | 0.40–2.13 | 0.859 | 0.87 | 0.37–2.02 | 0.746 |
| Widowed | 1.16 | 0.70–1.92 | 0.564 | 1.42 | 0.83–2.44 | 0.202 |
| Cohabiting | 0.76 | 0.60–0.98 | 0.032 | 0.90 | 0.69–1.17 | 0.429 |
| **Poverty status (ref: not poor)** |  |  |  |  |  |  |
| Poor | 0.72 | 0.56–0.94 | 0.016 | 0.87 | 0.64–1.18 | 0.367 |
| Extreme poverty | 0.46 | 0.33–0.64 | <0.001 | 0.66 | 0.46–0.93 | 0.017 |
|  | cPR | 95% CI | p | aPR | 95% CI | p |
| **Area of residence (ref: urban)** |  |  |  |  |  |  |
| Rural | 0.38 | 0.29–0.51 | <0.001 | 0.54 | 0.39–0.74 | <0.001 |
| **Natural region (ref: Metropolitan Lima)** |  |  |  |  |  |  |
| Coast | 0.75 | 0.58–0.96 | 0.023 | 0.81 | 0.63–1.04 | 0.102 |
| Highlands | 0.62 | 0.47–0.83 | 0.001 | 0.81 | 0.60–1.09 | 0.163 |
| Amazonian region | 0.46 | 0.31–0.69 | <0.001 | 0.62 | 0.40–0.95 | 0.027 |
| **Macroregion (ref: Metropolitan Lima)** |  |  |  |  |  |  |
| North | 0.63 | 0.45–0.86 | 0.004 | — | — | — |
| Central | 0.72 | 0.55–0.93 | 0.013 | — | — | — |
| South | 0.64 | 0.49–0.84 | 0.001 | — | — | — |
| Oriente | 0.49 | 0.30–0.81 | 0.005 | — | — | — |

**Panel C. Adults meeting survey-based criteria for ≥1 12-month mental disorder: self-perceived need**
| Variable | Crude |  |  | Adjusted |  |  |
| --- | --- | --- | --- | --- | --- | --- |
|  | cPR | 95% CI | p | aPR | 95% CI | p |
| Age (continuous) | 0.996 | 0.99–1.01 | 0.422 | 0.997 | 0.99–1.01 | 0.573 |
| **Sex** |  |  |  |  |  |  |
| Female vs male | 1.49 | 1.15–1.94 | 0.003 | 1.53 | 1.19–1.98 | 0.001 |
| **Educational attainment (ref: complete secondary)** |  |  |  |  |  |  |
| No education | 0.79 | 0.37–1.67* | 0.534 | 0.86 | 0.40–1.86* | 0.709 |
| Less than complete secondary | 0.89 | 0.69–1.14 | 0.342 | 1.04 | 0.80–1.35 | 0.762 |
| Higher education | 1.38 | 1.06–1.79 | 0.017 | 1.43 | 1.11–1.85 | 0.006 |
| **Marital status (ref: single)** |  |  |  |  |  |  |
| Married | 0.80 | 0.57–1.13 | 0.205 | 0.75 | 0.54–1.04 | 0.088 |
| Separated | 0.88 | 0.64–1.23 | 0.459 | 0.82 | 0.60–1.12 | 0.208 |
| Divorced | 2.00 | 1.34–2.97* | <0.001 | 1.27 | 0.88–1.85* | 0.200 |
| Widowed | 1.23 | 0.79–1.89 | 0.360 | 1.11 | 0.72–1.72 | 0.630 |
| Cohabiting | 0.97 | 0.74–1.27 | 0.842 | 0.94 | 0.72–1.23 | 0.654 |
| **Poverty status (ref: not poor)** |  |  |  |  |  |  |
| Poor | 0.79 | 0.63–1.00 | 0.050 | 0.86 | 0.68–1.10 | 0.233 |
| Extreme poverty | 0.70 | 0.50–0.99 | 0.043 | 0.82 | 0.57–1.16 | 0.264 |
| **Area of residence (ref: urban)** |  |  |  |  |  |  |
| Rural | 0.60 | 0.45–0.81 | <0.001 | 0.78 | 0.56–1.07 | 0.117 |
| **Natural region (ref: Metropolitan Lima)** |  |  |  |  |  |  |
| Coast | 0.94 | 0.72–1.24 | 0.678 | 0.999 | 0.77–1.30 | 0.995 |
| Highlands | 0.81 | 0.60–1.09 | 0.171 | 0.95 | 0.72–1.25 | 0.694 |
| Amazonian region | 0.75 | 0.52–1.07 | 0.116 | 0.87 | 0.61–1.25 | 0.460 |
| **Macroregion (ref: Metropolitan Lima)** |  |  |  |  |  |  |
| North | 0.94 | 0.68–1.28 | 0.689 | — | — | — |
| Central | 0.94 | 0.71–1.24 | 0.637 | — | — | — |
| South | 0.75 | 0.56–1.02 | 0.069 | — | — | — |
| Oriente | 0.67 | 0.44–1.02 | 0.061 | — | — | — |

**Panel D. Adults meeting survey-based criteria for ≥1 12-month mental disorder: need identified by others**
| Variable | Crude |  |  | Adjusted |  |  |
| --- | --- | --- | --- | --- | --- | --- |
|  | cPR | 95% CI | p | aPR | 95% CI | p |
| Age (continuous) | 0.99 | 0.97–1.00 | 0.027 | 0.99 | 0.98–1.00 | 0.121 |
| **Sex** |  |  |  |  |  |  |
| Female vs male | 1.10 | 0.80–1.51 | 0.569 | 1.08 | 0.78–1.50 | 0.640 |
| **Educational attainment (ref: complete secondary)** |  |  |  |  |  |  |
| No education | 0.66 | 0.23–1.93* | 0.449 | 0.90 | 0.29–2.81* | 0.857 |
| Less than complete secondary | 0.64 | 0.46–0.89 | 0.008 | 0.87 | 0.61–1.24 | 0.429 |
| Higher education | 0.99 | 0.66–1.50 | 0.981 | 1.09 | 0.74–1.62 | 0.657 |
|  | cPR | 95% CI | p | aPR | 95% CI | p |
| **Marital status (ref: single)** |  |  |  |  |  |  |
| Married | 0.56 | 0.31–1.01 | 0.054 | 0.70 | 0.38–1.29 | 0.253 |
| Separated | 1.14 | 0.76–1.72 | 0.526 | 1.37 | 0.91–2.06 | 0.133 |
| Divorced | 0.33 | 0.06–1.82* | 0.204 | 0.33 | 0.05–1.96* | 0.221 |
| Widowed | 1.36 | 0.80–2.32 | 0.250 | 1.91 | 1.08–3.36 | 0.026 |
| Cohabiting | 0.93 | 0.65–1.33 | 0.705 | 1.15 | 0.81–1.63 | 0.425 |
| **Poverty status (ref: not poor)** |  |  |  |  |  |  |
| Poor | 0.66 | 0.47–0.92 | 0.014 | 0.74 | 0.52–1.04 | 0.081 |
| Extreme poverty | 0.45 | 0.28–0.73 | 0.001 | 0.51 | 0.31–0.83 | 0.006 |
| **Area of residence (ref: urban)** |  |  |  |  |  |  |
| Rural | 0.44 | 0.28–0.67 | <0.001 | 0.66 | 0.42–1.05 | 0.081 |
| **Natural region (ref: Metropolitan Lima)** |  |  |  |  |  |  |
| Coast | 0.85 | 0.59–1.23 | 0.389 | 0.91 | 0.63–1.31 | 0.617 |
| Highlands | 0.79 | 0.51–1.23 | 0.294 | 0.91 | 0.60–1.38 | 0.662 |
| Amazonian region | 0.52 | 0.32–0.87 | 0.012 | 0.65 | 0.39–1.08 | 0.096 |
| **Macroregion (ref: Metropolitan Lima)** |  |  |  |  |  |  |
| North | 1.05 | 0.68–1.61 | 0.834 | — | — | — |
| Central | 0.71 | 0.48–1.04 | 0.082 | — | — | — |
| South | 0.58 | 0.39–0.88 | 0.009 | — | — | — |
| Oriente | 0.62 | 0.37–1.06 | 0.082 | — | — | — |

cPR, crude prevalence ratio; aPR, adjusted prevalence ratio; CI, confidence interval. p values tested individual coefficients and accounted for the complex survey design. The derived measure retained valid direct responses among participants without annual contact and used SR111 and SR113 among participants with annual contact. Adjusted models included age, sex, educational attainment, marital status, poverty status, area of residence, and natural region; macroregion is presented only in crude analyses. \* Category with unweighted n < 30 or fewer than five events or non-events; interpret cautiously. ENSM, National Mental Health Survey.

### 3.6 Use of mental health services and perceived quality of care

Annual psychological service use was 3.7% (95% CI 3.2–4.3; 512/13,124), while psychiatric service use was 1.1% (95% CI 0.8–1.5; 120/13,170). Lifetime use of a mental health telephone line was 0.5% (95% CI 0.3–0.7) (Table 6).

**Table 6.** Mental health service use: annual psychiatry and psychology care and lifetime telephone-line use, ENSM Peru 2022.

| Variable | n/N | % | 95% CI |
| --- | --- | --- | --- |
| Annual psychiatry service use | 120/13,170 | 1.1 | 0.8–1.5 |
| Annual psychology service use | 512/13,124 | 3.7 | 3.2–4.3 |
| **Satisfaction with psychiatry care (users, n=120)** |  |  |  |
| Very dissatisfied | 2 | 1.6 | 0.3–7.4 |
| Dissatisfied | 9 | 5.7 | 2.2–13.9 |
| Neither satisfied nor dissatisfied | 19 | 21.2 | 10.4–38.6 |
| Satisfied | 64 | 57.4 | 41.6–71.8 |
| Very satisfied | 26 | 14.1 | 7.8–24.1 |
| **Perceived helpfulness of psychiatry care (users, n=120)** |  |  |  |
| Not at all | 6 | 3.6 | 1.3–9.6 |
| A little | 13 | 16.8 | 7.2–34.5 |
| Moderately | 39 | 27.4 | 16.8–41.3 |
| A lot | 62 | 52.2 | 36.5–67.4 |
| **Satisfaction with psychology care (evaluable users, n=510)** |  |  |  |
| Very dissatisfied | 9 | 1.4 | 0.7–3.1 |
| Dissatisfied | 29 | 6.4 | 3.9–10.3 |
| Neither satisfied nor dissatisfied | 56 | 15.9 | 10.9–22.6 |
| Satisfied | 283 | 51.5 | 44.6–58.3 |
| Very satisfied | 133 | 24.9 | 19.2–31.6 |
| **Perceived helpfulness of psychology care (evaluable users, n=511)** |  |  |  |
| Not at all | 28 | 5.9 | 3.2–10.4 |
| A little | 75 | 14.7 | 11.0–19.5 |
| Moderately | 118 | 27.3 | 21.5–34.0 |
| A lot | 290 | 52.0 | 45.1–58.9 |
| Lifetime mental health telephone-line use | 64/13,205 | 0.5 | 0.3–0.7 |
| **Time since last telephone-line use** |  |  |  |
| Past month | 7 | 15.7 | 3.6–47.8 |
| 2–6 months | 10 | 23.6 | 8.5–50.6 |
| 7–12 months | 8 | 14.8 | 5.0–36.4 |
| More than 1 year | 37 | 43.3 | 25.2–63.4 |
| Do not know | 2 | 2.6 | 0.6–10.9 |

For annual service-use rows, n/N denotes users/participants with an evaluable outcome: 120/13,170 for psychiatry and 512/13,124 for psychology. Survey commands used 13,163 and 13,117 observations, respectively, because seven records lacked complete design information. Satisfaction and perceived-helpfulness estimates were restricted to evaluable service users. Telephone-line use denotes lifetime use; time-since-last-use categories were calculated among respondents reporting use. Percentages and 95% CIs account for the complex survey design. CI, confidence interval; ENSM, National Mental Health Survey.

Among service users, most respondents reported being satisfied or very satisfied and rated care as at least moderately helpful. These distributions were estimated only among 120 psychiatric users and approximately 510 psychological users and do not represent the experience of adults without access (Table 6).

#### i. Exploratory crude factors associated with access and user-reported experience

Crude associations with service access are reported in Supplementary Tables S2A and S2B. Psychiatric and psychological access were lower in several socioeconomically and geographically disadvantaged groups, including adults in extreme poverty and rural residents; psychological access was higher among women.

Associations with satisfaction and perceived helpfulness were heterogeneous and were based on small service-user subpopulations, particularly for psychiatry. These analyses involved multiple comparisons and were treated as exploratory (Supplementary Table S2B).

Telephone-line use was lower among rural residents and residents of several regions outside Metropolitan Lima, although this outcome reflected lifetime rather than 12-month use (Supplementary Table S2B).

## 4. DISCUSSION

### 4.1 Principal findings

In this nationally representative analysis of Peruvian adults, the mental health contact gap was 61.0% among adults meeting survey-based criteria for a lifetime disorder and 84.5% among those with a 12-month disorder. Rural estimates exceeded urban estimates in both periods; after adjustment, poverty and rural residence were associated with the lifetime contact gap, while extreme poverty was associated with the 12-month gap. Perceived need was also unequally distributed, although adjusted patterns differed between the general population and adults meeting survey-based criteria for one or more 12-month mental disorders. Psychological and psychiatric service use remained low, whereas satisfaction and perceived helpfulness were relatively high among users. These results describe social and territorial patterning of reported contact and should not be interpreted as measures of adequate or effective treatment or as an estimate of the effect of mental health reform.

### 4.2 Strengths and limitations

Before interpreting these findings, several strengths and limitations should be acknowledged. The ENSM 2022 used a national probabilistic multistage design, standardized diagnostic assessment, calibrated survey weights, and explicit criteria for statistical precision. The large adult sample and coverage of all 25 political regions allowed national, regional, and urban–rural estimates, while sensitivity analyses clarified how the definition of contact affected the main estimates.

Selection bias cannot be excluded. Household non-response was not fully documented in the available materials, and the exclusion of people with hearing, cognitive, neurological, or communication difficulties may have underrepresented adults with greater clinical or functional impairment. The survey also excluded institutionalized populations, including people in hospitals, long-term care facilities, prisons, and shelters. Because these groups may differ in both mental health needs and contact with services, the direction of the resulting bias is uncertain and the estimates should not be generalized to them.

Service use, perceived need, satisfaction, and helpfulness were self-reported and may have been affected by recall, non-disclosure, stigma, or social desirability. The perceived-need outcomes were analytically derived by routing direct variables with SR111 and SR113 rather than by applying an official ENSM equivalence, and classification error is possible if these items capture nonidentical constructs. The main prevalence estimates were stable when observations with missing annual contact status were excluded, but two borderline subgroup coefficients crossed the conventional p = 0.05 threshold and were therefore not treated as robust. Lifetime contact is particularly vulnerable to recall error and relied on a global indicator that could not be fully reconstructed from provider-specific items; the 12-month broad-contact gap was therefore treated as the primary estimand. Satisfaction and helpfulness were observed only among people who reached services and may reflect selection into care, expectations, or continued engagement among those with more favourable experiences. They cannot be interpreted as objective measures of treatment quality or system performance.

Mental disorders were identified using survey algorithms derived from the World Health Organization Composite International Diagnostic Interview rather than clinical evaluation. Diagnostic misclassification is therefore possible, particularly for less frequent conditions with imprecise estimates. In addition, the available secondary dataset did not provide reproducible general measures of severity, functioning, recurrence, episode duration, or persistence. Recent diagnostic activity and the five-domain count were used only as exploratory indicators of clinical complexity and should not be interpreted as persistence, a complete count of comorbidity, or comprehensive clinical need.

Finally, the cross-sectional design precludes temporal or causal interpretation. Poverty, residence, region, perceived need, and service use may influence one another and may share unmeasured determinants such as local service availability, distance, insurance, health literacy, or stigma. The adjusted prevalence ratios were intended to describe model-conditional patterning, not total causal effects; interpreting every covariate coefficient causally would reproduce the ‘Table 2 fallacy’ (36,37). Data collection in late 2022 may also have reflected pandemic-related changes, although this survey cannot separate those influences from longer-standing barriers.

### 4.3 Interpretation

The magnitude of the contact gaps is broadly compatible with World Mental Health Survey analyses and prior evidence from Latin America and Peru, which have reported large treatment gaps in upper-middle-income and lower-middle-income settings (1,8,9,13). Comparisons require caution because studies differ in included disorders, diagnostic instruments, reference periods, provider definitions, and whether the outcome represents any contact or minimally adequate treatment. In the present study, the lifetime and 12-month estimates are separate cross-sectional estimands; their difference does not demonstrate delayed help-seeking, eventual access, or an individual trajectory from disorder onset to care.

Earlier Peruvian evidence based on the World Mental Health Survey collected in 2005 also documented substantial unmet need, although it cannot be treated as a directly comparable baseline (13). The surveys differ in calendar period, diagnostic algorithms, included disorders, provider definitions, and operationalization of contact. In addition, the present lifetime estimate relied on the global survey indicator, whereas the annual estimate could be reconstructed from named provider categories. A valid assessment of change over time would require harmonized variables, common eligibility rules, equivalent survey design specifications, and an explicit comparison model. The current findings therefore provide a contemporary population description rather than evidence that contact gaps persisted, improved, or worsened relative to the earlier survey.

The timing of the survey provides additional context but does not resolve this temporal question. Data were collected in November and December 2022, after the most acute phase of COVID-19- related service disruption. Villarreal-Zegarra et al. reported that Community Mental Health Centers experienced an initial decline in users and appointments in March 2020, followed by recovery in service volume during the pandemic (16). More recent nationwide data indicate that recovery and decentralization were not uniform across diagnostic groups: between 2018 and 2024, service utilization increased for non-psychotic mental disorders but decreased for psychosis, despite a shift toward primary and regional facilities (22). However, recovery in aggregate volume does not establish recovery in equitable coverage, continuity, treatment adequacy, or perceived accessibility. Pandemic-related changes in help-seeking, household income, mobility, and the availability of in-person or remote services may still have influenced the 2022 estimates. Because the ENSM provides a single cross-sectional measurement, these influences cannot be separated from longer-standing structural and demand-side barriers.

The association of poverty and rural residence with contact gaps is consistent with evidence that cost, distance, transportation, workforce distribution, and uneven service availability constrain access (19,23,24). In Peru, recent qualitative evidence from three northern provinces suggests that improved geographic proximity to community-based services may coexist with fragmented referral pathways, workforce instability, limitations in follow-up, stigma, economic constraints, and tensions between cultural and biomedical understandings of mental health (19). However, these mechanisms were not measured directly. The observed prevalence associations identify groups with lower contact but do not show which barrier caused the difference or whether service expansion changed it.

These overlapping constraints are likely to operate at several levels. At the individual and household levels, transportation costs, time away from paid or unpaid work, health literacy, stigma, and competing needs may affect whether care is sought. At the service level, workforce availability, referral pathways, opening hours, medication supply, and continuity between primary care and specialized services may shape whether an initial contact occurs and is sustained. Territorial factors may further modify these barriers through distance, language, connectivity, and the distribution of Community Mental Health Centres. The ENSM did not directly measure most of these mechanisms, and residence or poverty should not be treated as causal substitutes for them. Nevertheless, their consistent descriptive patterning identifies populations for whom more detailed assessment of the pathway to care is warranted.

The adjusted models should be read as a description of these jointly distributed inequalities rather than as an attempt to isolate a single determinant. Education, poverty, marital status, rural residence, and natural region may share antecedents and may also lie on overlapping pathways to service contact. Mutual adjustment can therefore attenuate an estimate without showing that the corresponding barrier is unimportant. Conversely, a remaining prevalence ratio does not establish an independent causal effect, because severity, comorbidity, insurance, travel time, service availability, language, social support, and previous experiences with care were not represented comprehensively in these models. The estimates are most informative when considered together with the absolute prevalences: rural adults had visibly larger contact gaps in the descriptive analysis, while the adjusted annual association was smaller and its confidence interval included the null. This combination is compatible with partial statistical explanation by measured covariates, limited precision, residual confounding, or several of these factors at once. It should not be summarized as evidence that residence ceased to matter. For monitoring, the practical implication is to report both absolute gaps and adjusted relative measures, retain uncertainty intervals, and avoid converting conventional significance thresholds into a binary judgment about equity. Future analyses could evaluate prespecified interactions and incorporate contextual measures of service supply, but they would require adequate sample size within survey domains and a causal framework defined before model fitting.

Perceived need for care was substantially more frequent among adults meeting survey-based criteria for one or more 12-month mental disorders than in the general population. Within this group, self-perceived need was reported by 37.3% (95% CI 33.4–41.4), while need identified by others was reported for 25.6% (95% CI 21.8–29.8). Adjusted self-perceived need was higher among women and adults with higher education, whereas need identified by others was less frequent among adults in extreme poverty. Crude urban–rural differences in self-perceived need attenuated after adjustment, illustrating that descriptive geographic contrasts and model-conditional associations provide complementary information. For need identified by others in this subgroup, the adjusted rural and Amazonian-region estimates included the null; the rural estimate was also sensitive to the handling of missing contact status and was not interpreted as robust. These patterns may reflect differences in mental health literacy, stigma, competing priorities, previous experience with services, or culturally specific interpretations of distress, although the ENSM 2022 did not directly measure these mechanisms (12,20,21,25,26).

Perceived need should also be distinguished from clinical need. Recent World Mental Health Survey analyses further separate problem recognition from the belief that professional help is required, components that may have different predictors (20). The survey-based diagnostic algorithms identify respondents who met symptom criteria, whereas self-perceived need reflects whether respondents recognized or articulated a reason to seek professional care; need identified by others reflects a different social process. Neither measure determines whether treatment was clinically indicated, wanted, available, or adequate. This distinction is particularly relevant because the adjusted associations were not identical across the general population and the subgroup meeting survey-based criteria for one or more 12-month mental disorders. Analyses in the general population describe how perceived need is distributed across all adults, while analyses restricted to respondents meeting the survey algorithm for one or more 12-month mental disorders address recognition conditional on the survey algorithm. Presenting both denominators prevents the lower population prevalence from being mistaken for low recognition among adults meeting the survey algorithm.

Exploratory analyses found smaller contact gaps among respondents with recent diagnostic activity and among those with two or more available diagnostic domains. Greater symptom visibility, family recognition, or opportunities for detection may contribute, although severity and functioning were unavailable. Contact gaps also varied by diagnostic group, with high estimates for alcohol- and other substance-use disorders, a pattern compatible with evidence of low treatment coverage and perceived need for these conditions (1,27). Conversely, lower point estimates for psychotic disorders may reflect greater clinical visibility, but sparse data and wide confidence intervals preclude firm comparisons (13,28). Recent Peruvian service-use data also caution against interpreting psychosis as a uniformly better-covered group, because utilization for psychosis declined between 2018 and 2024 while utilization for non-psychotic mental disorders increased (22).

Disorder-specific estimates require the same caution. Alcohol- and substance-use disorders may involve stigma, normalization of use, lower perceived need, and limited integration of addiction treatment into primary and community mental health services (27). Psychotic symptoms, in contrast, may be more visible to relatives or services and may prompt contact even when the affected person does not identify a need. However, the apparent differences between diagnostic groups may also reflect variation in algorithm performance, symptom severity, provider pathways, and sparse survey domains. Several confidence intervals were wide or non-informative, especially in rural or regional strata. These results are therefore best viewed as descriptive signals for future study, not as a ranking of which disorders receive adequate care.

Annual use of psychological and psychiatric services was low in the adult population, whereas reported satisfaction and helpfulness were relatively high among users. Cross-national World Mental Health Survey data similarly show generally high patient-reported satisfaction and helpfulness among people who reach care, with variation by provider type and patient characteristics (29). These observations are not contradictory because they refer to different denominators: coverage concerns the population, while experience measures are conditional on reaching a service. People who obtain care may differ in socioeconomic position, proximity, health literacy, symptom recognition, expectations, and prior experiences (30–32). High satisfaction among this selected group can therefore coexist with restricted population coverage and should not be used as evidence of adequate, continuous, or effective care. Moreover, satisfaction measures can be sensitive to survey design and framing and should complement rather than substitute for more objective indicators of quality (33).

The exploratory associations with user-reported experience reinforce this denominator problem. Some crude estimates suggested greater satisfaction or helpfulness in disadvantaged groups, but the analyses involved multiple comparisons and small service-user samples, particularly the 120 psychiatric users. Such patterns may reflect real differences in experience, but they may also arise from imprecision, expectations, the type of service reached, or selective continuation among people who perceived benefit. The data did not measure guideline-concordant treatment, dose, duration, therapeutic continuity, clinical improvement, or recovery. Accordingly, satisfaction and helpfulness should complement rather than replace measures of coverage, timeliness, continuity, adequacy, functioning, and quality of life.

The ENSM 2022 was conducted after substantial expansion of community-based mental health infrastructure, but the present design was not an evaluation of that reform. The findings show that high contact gaps and social inequalities coexisted with the expanded network; they do not establish what the gaps would have been without the reform or whether they increased or decreased because of it. Monitoring should therefore distinguish service volume from population coverage and incorporate equity, perceived need, continuity, adequacy, functioning, and recovery-oriented outcomes (14,15,34,35).

This distinction matters for the interpretation of reform. Counts of centers, consultations, or reported contacts describe service production, while a population contact gap describes the proportion of adults meeting a survey algorithm who did not report contact during a defined period. Neither measure alone captures whether care was timely, continuous, culturally responsive, or clinically adequate. This distinction also parallels contemporary coverage frameworks that separate contact from minimally adequate and effective treatment (9,10). A stronger evaluation framework would combine repeated population surveys with routine service data and indicators of need, functioning, continuity, treatment adequacy, and equity. It would also specify a comparison strategy capable of separating secular trends and pandemic effects from implementation of the community-based model. Until such evidence is available, the present findings should be used to identify monitoring priorities rather than to judge the causal success or failure of the reform.

### 4.4 Implications

The results support closer surveillance of contact gaps among adults in poverty and rural or geographically underserved areas. Potential responses include strengthening referral pathways between primary care and Community Mental Health Centers, improving culturally adapted mental health literacy and community engagement, and reducing indirect costs related to distance and time (19). These are programmatic implications rather than effects demonstrated by the study; specific strategies should be evaluated for feasibility, acceptability, coverage, equity, and sustainability before broad implementation.

Further research should use longitudinal designs or linked service records to examine the timing, continuity, and adequacy of care; incorporate validated measures of severity and functioning; and evaluate whether reforms change contact gaps over time. Research should also clarify why perceived need and service contact differ across social and territorial groups and should examine experiences among people who never reach services, rather than relying only on satisfaction among users.

## 5. CONCLUSIONS

Mental health contact gaps were high among Peruvian adults meeting survey-based criteria for mental disorders, particularly for 12-month disorders, and were patterned by poverty and rural residence. Perceived need and service contact were also unequally distributed, while satisfaction and helpfulness described only the selected group who reached care. These findings support monitoring population contact, equity, continuity, and adequacy alongside service expansion, but they do not establish individual care trajectories, effective treatment, or the causal impact of mental health reform.

## 6. OTHER INFORMATION

### Ethics

The secondary-analysis project entitled “Brecha de Atención en Salud Mental en Perú durante el año 2022: un estudio de bases secundarias” was officially registered by the Instituto Nacional de Salud Mental “Honorio Delgado – Hideyo Noguchi” under OEAIDE pre-project code 1010-2025 (approved on November 26, 2025) and institutional code INSM 614-2025 (officialized on December 11, 2025). On December 11, 2025, the Institutional Research Ethics Committee of the same institute issued an exemption from protocol review for the secondary-analysis project (Constancia de Exención de Revisión de un Protocolo de Investigación No. 020-2025-CIEI- INSM “HD-HN”). The certificate states that the exemption covers the research project and its annexes and is valid for one year from the date of issuance. This secondary analysis did not recruit participants or collect new participant data.

### Data availability

The ENSM 2022 database was provided by OEAIDE for use in this institutionally authorized secondary-analysis project.

### Author contributions

Paulo Ruiz-Grosso: Conceptualization, Methodology, Investigation, Data curation, Formal analysis, Software, Validation, Visualization, Project administration, Writing – original draft, Writing – review & editing. Luis Enrique Macedo Orrego: Conceptualization, Methodology, Writing – original draft, Writing – review & editing. María Teresa Rivera Encinas, María Soledad Carazas Vera, Diego Mauricio Rodríguez Vargas, and Alejandra Arosemena Aliaga: Writing – original draft, Writing – review & editing. Abel Ampelio II Sagástegui Soto and Sonia Zevallos Bustamante: Project administration, Supervision, Writing – original draft, Writing – review & editing. All authors approved the final version and accept responsibility for the work.

### Use of generative artificial intelligence

During manuscript preparation, the authors used OpenAI ChatGPT and Codex (OpenAI, San Francisco, California; used from August 4 to 19, 2026) to assist with language editing, structural revision, organization of analytical documentation, and consistency checking. The AI-assisted review process was structured using an author-developed quality-assurance workflow. All AI-assisted content was critically reviewed and revised by the authors, who take full responsibility for the accuracy, integrity, and final content of the manuscript.

