## Supplemental Tables for "Mental health contact gaps among adults in Peru: a cross-sectional analysis of a nationally representative survey"

**Contents**

Supplementary Table S1. Sensitivity of contact-gap estimates to alternative contact definitions.

Supplementary Table S2A. Crude exploratory associations with annual psychiatry and psychology service use, with model and category counts.

Supplementary Table S2B. Crude exploratory associations with annual service use, user-reported experience, and telephone-line use.

Supplementary Table S3. Exploratory indicators of clinical complexity and annual contact gaps.

Supplementary Table S4. Analytic variable dictionary.

*All estimates account for the complex survey design of the ENSM Peru 2022 (raked_weight_trimmed as the probability weight, ccluster as the first-stage primary sampling unit, and departamentos_si_callao as the stratum, with singleton strata centred). Contact and perceived-need measures describe reported contact and recognition of need; they do not establish that care was adequate, continuous, or effective.*

**Supplementary Table S1. Sensitivity of contact-gap estimates to alternative contact definitions, ENSM Peru 2022**

*Population: adults meeting survey-based criteria for one or more mental disorders during the corresponding period*

**Panel A. Lifetime contact gap**

| **Contact definition** | **Included sources** | **Evaluable n** | **n without contact** | **Contact gap, %** | **95% CI** | **Interpretation** |
| --- | --- | --- | --- | --- | --- | --- |
| Broad contact: any included source | Psychiatrist; psychologist; general practitioner/other specialist; social worker; counsellor/community agent; other mental health professional; other health professional; religious/spiritual adviser; traditional healer | 3927 | 2422 | 61.0 | 58.4–63.7 | Includes health and non-health sources; it does not necessarily indicate formal treatment. |
| Specialists: psychiatrist or psychologist | Psychiatrist; psychologist | 3926 | 2863 | 72.5 | 70.1–74.9 | More restrictive definition; by construction, it yields the largest contact gap. |
| Specialists plus physician | Psychiatrist; psychologist; general practitioner or other specialist | 3926 | 2713 | 68.7 | 66.2–71.2 | Adding physicians reduces the gap relative to considering only psychiatry or psychology. |
| Expanded health sector | Psychiatrist; psychologist; general practitioner/other specialist; other health professional | 3926 | 2700 | 68.5 | 66.0–71.0 | Adding other health professionals changes the estimate little relative to specialists plus physician. |

**Panel B. Twelve-month contact gap**

| **Contact definition** | **Included sources** | **Evaluable n** | **n without contact** | **Contact gap, %** | **95% CI** | **Interpretation** |
| --- | --- | --- | --- | --- | --- | --- |
| Broad contact: any included source | Psychiatrist; psychologist; general practitioner/other specialist; social worker; counsellor/community agent; other mental health professional; other health professional; religious/spiritual adviser; traditional healer | 1649 | 1382 | 84.5 | 81.5–87.6 | Primary definition used in the manuscript; includes health and non-health sources and does not necessarily indicate formal treatment. |
| Specialists: psychiatrist or psychologist | Psychiatrist; psychologist | 1648 | 1452 | 89.0 | 86.4–91.6 | More restrictive definition; by construction, it yields the largest contact gap. |
| Specialists plus physician | Psychiatrist; psychologist; general practitioner or other specialist | 1649 | 1418 | 87.5 | 84.8–90.2 | Adding physicians reduces the gap relative to considering only psychiatry or psychology. |
| Expanded health sector | Psychiatrist; psychologist; general practitioner/other specialist; other health professional | 1649 | 1416 | 87.4 | 84.7–90.1 | Adding other health professionals changes the estimate little relative to specialists plus physician. |

Notes. Results were weighted for the complex ENSM Peru 2022 survey design; evaluable n and n without contact are unweighted counts. The contact gap denotes absence of contact. CI, confidence interval; ENSM, National Mental Health Survey.

The broad 12-month definition uses mhs_anual and includes health and non-health sources; it should therefore be described as contact with any source of mental health-related care rather than formal treatment.

The equivalence of mhs_vida with the provider-specific indicators available in the analytic files could not be fully documented. Lifetime estimates should therefore be interpreted descriptively; the 12-month broad-contact definition is the primary definition used in the manuscript.

In an additional missing-data sensitivity analysis for the expanded health-sector definition at 12 months, excluding zero values with any missing provider indicator yielded a contact gap of 87.1% (95% CI 84.3–89.9; evaluable n=1,621), which was similar to the primary estimate.

The two 12-month rows above were recovered from the reproducible file S1_resultados_exactos.csv (values already stored in v06 but not displayed in the sheet) and re-verified before use.

**Supplementary Table S2A. Crude exploratory associations with annual psychiatry and psychology service use, ENSM Peru 2022**

**Panel A. Annual psychiatry service use**

| **Variable** | **Category** | **Model n** | **Users** | **Category n** | **Category users** | **cPR** | **95% CI** | **p** | **Global p** | **Note** |
| --- | --- | --- | --- | --- | --- | --- | --- | --- | --- | --- |
| Age (continuous) | Per year | 13163 | 120 | 13163 | 120 | 0.997 | 0.97–1.03 | 0.845 | 0.845 |  |
| Sex (ref: male) | Female | 13141 | 119 | 8609 | 90 | 0.996 | 0.52–1.91 | 0.991 | 0.991 |  |
| Educational attainment (ref: complete secondary) | No education | 13145 | 120 | 278 | 0 | NE | NE | NE | NE | NE: model did not converge |
|  | Less than complete secondary | 13145 | 120 | 3951 | 19 | NE | NE | NE | NE | NE: model did not converge |
|  | Higher education | 13145 | 120 | 2900 | 47 | NE | NE | NE | NE | NE: model did not converge |
| Marital status (ref: single) | Married | 13085 | 120 | 3275 | 34 | 1.07 | 0.47–2.43 | 0.864 | <0.001 |  |
|  | Separated | 13085 | 120 | 1303 | 14 | 1.12 | 0.38–3.32 | 0.840 |  |  |
|  | Divorced | 13085 | 120 | 141 | 4 | 3.62 | 0.99–13.25 | 0.052 |  | * |
|  | Widowed | 13085 | 120 | 270 | 3 | 1.87 | 0.37–9.43 | 0.447 |  | * |
|  | Cohabiting | 13085 | 120 | 4306 | 14 | 0.22 | 0.10–0.50 | <0.001 |  |  |
| Poverty status (ref: not poor) | Poor | 13163 | 120 | 4114 | 25 | 0.42 | 0.22–0.82 | 0.011 | <0.001 |  |
|  | Extreme poverty | 13163 | 120 | 1513 | 4 | 0.04 | 0.01–0.14 | <0.001 |  | * |
| Area of residence (ref: urban) | Rural | 13150 | 120 | 2841 | 8 | 0.07 | 0.02–0.18 | <0.001 | <0.001 | * |
| Natural region (ref: Metropolitan Lima) | Coast | 13147 | 120 | 4218 | 40 | 0.43 | 0.22–0.84 | 0.013 | 0.007 |  |
|  | Highlands | 13147 | 120 | 5098 | 38 | 0.37 | 0.18–0.74 | 0.005 |  |  |
|  | Amazonian region | 13147 | 120 | 1955 | 12 | 0.26 | 0.11–0.61 | 0.002 |  |  |
| Macroregion (sensitivity analysis; ref: Metropolitan Lima) | North | 13153 | 120 | 2910 | 22 | 0.35 | 0.17–0.73 | 0.005 | 0.003 |  |
|  | Central | 13153 | 120 | 3624 | 25 | 0.27 | 0.13–0.56 | <0.001 |  |  |
|  | South | 13153 | 120 | 3597 | 36 | 0.58 | 0.29–1.18 | 0.132 |  |  |
|  | Oriente | 13153 | 120 | 1146 | 7 | 0.28 | 0.10–0.76 | 0.013 |  | * |

**Panel B. Annual psychology service use**

| **Variable** | **Category** | **Model n** | **Users** | **Category n** | **Category users** | **cPR** | **95% CI** | **p** | **Global p** | **Note** |
| --- | --- | --- | --- | --- | --- | --- | --- | --- | --- | --- |
| Age (continuous) | Per year | 13117 | 512 | 13117 | 512 | 0.98 | 0.97–0.99 | <0.001 | <0.001 |  |
| Sex (ref: male) | Female | 13094 | 510 | 8582 | 403 | 2.03 | 1.47–2.80 | <0.001 | <0.001 |  |
| Educational attainment (ref: complete secondary) | No education | 13100 | 512 | 277 | 2 | 0.03 | 0.01–0.18 | <0.001 | <0.001 | * |
|  | Less than complete secondary | 13100 | 512 | 3948 | 116 | 0.62 | 0.45–0.84 | 0.002 |  |  |
|  | Higher education | 13100 | 512 | 2889 | 151 | 1.32 | 0.97–1.79 | 0.081 |  |  |
| Marital status (ref: single) | Married | 13038 | 511 | 3270 | 113 | 0.61 | 0.42–0.87 | 0.006 | 0.004 |  |
|  | Separated | 13038 | 511 | 1299 | 72 | 1.14 | 0.72–1.79 | 0.576 |  |  |
|  | Divorced | 13038 | 511 | 140 | 7 | 1.73 | 0.68–4.36 | 0.247 |  | * |
|  | Widowed | 13038 | 511 | 266 | 7 | 0.75 | 0.32–1.74 | 0.503 |  | * |
|  | Cohabiting | 13038 | 511 | 4289 | 139 | 0.65 | 0.47–0.90 | 0.009 |  |  |
| Poverty status (ref: not poor) | Poor | 13117 | 512 | 4104 | 131 | 0.62 | 0.47–0.81 | <0.001 | <0.001 |  |
|  | Extreme poverty | 13117 | 512 | 1508 | 27 | 0.26 | 0.16–0.45 | <0.001 |  |  |
| Area of residence (ref: urban) | Rural | 13105 | 511 | 2838 | 47 | 0.36 | 0.24–0.55 | <0.001 | <0.001 |  |
| Natural region (ref: Metropolitan Lima) | Coast | 13102 | 511 | 4205 | 209 | 1.40 | 0.95–2.05 | 0.087 | 0.004 |  |
|  | Highlands | 13102 | 511 | 5079 | 169 | 0.98 | 0.66–1.47 | 0.928 |  |  |
|  | Amazonian region | 13102 | 511 | 1949 | 61 | 0.70 | 0.44–1.11 | 0.127 |  |  |
| Macroregion (sensitivity analysis; ref: Metropolitan Lima) | North | 13108 | 512 | 2907 | 117 | 1.07 | 0.69–1.67 | 0.749 | 0.054 |  |
|  | Central | 13108 | 512 | 3611 | 147 | 1.16 | 0.78–1.72 | 0.456 |  |  |
|  | South | 13108 | 512 | 3579 | 141 | 1.28 | 0.84–1.93 | 0.249 |  |  |
|  | Oriente | 13108 | 512 | 1142 | 35 | 0.63 | 0.38–1.06 | 0.080 |  |  |

cPR, crude prevalence ratio; CI, confidence interval. Poisson models with a log link used survey weights and linearized standard errors for the complex ENSM Peru 2022 design. Analyses were exploratory and restricted to participants with the corresponding service-use indicator observed. Model n and Users describe the full model sample and events; Category n and Category users describe each predictor level. Global p values are from design-adjusted Wald tests for categorical predictors. * Fewer than 10 unweighted users in the category; interpret cautiously. The educational-attainment model for psychiatry did not converge because there were no users in the no-education category and is shown as NE. Macroregion was a sensitivity analysis. Psychiatry and psychology service-use indicators were missing for 670 and 716 participants, respectively; the reason for this item-level missingness could not be determined from the available documentation. ENSM, National Mental Health Survey; NE, not estimable.

**Supplementary Table S2B. Crude exploratory associations with annual service use, user-reported experience, and lifetime telephone-line use, ENSM Peru 2022**

**Panel A. Annual psychiatry service use**

| **Variable** | **cPR** | **95% CI** | **p** |
| --- | --- | --- | --- |
| Age (continuous) | 0.997 | 0.97–1.03 | 0.845 |
| **Sex** |  |  |  |
| Female vs male | 0.996 | 0.52–1.91 | 0.991 |
| **Educational attainment (ref: complete secondary)** |  |  |  |
| No education | NE | NE | NE |
| Less than complete secondary | NE | NE | NE |
| Higher education | NE | NE | NE |
| **Marital status (ref: single)** |  |  |  |
| Married | 1.07 | 0.47–2.43 | 0.864 |
| Separated | 1.12 | 0.38–3.32 | 0.840 |
| Divorced | 3.62 | 0.99–13.25 | 0.052 |
| Widowed | 1.87 | 0.37–9.43 | 0.447 |
| Cohabiting | 0.22 | 0.10–0.50 | <0.001 |
| **Poverty status (ref: not poor)** |  |  |  |
| Poor | 0.42 | 0.22–0.82 | 0.011 |
| Extreme poverty | 0.04 | 0.01–0.14 | <0.001 |
| **Area of residence (ref: urban)** |  |  |  |
| Rural | 0.07 | 0.02–0.18 | <0.001 |
| **Natural region (ref: Metropolitan Lima)** |  |  |  |
| Coast | 0.43 | 0.22–0.84 | 0.013 |
| Highlands | 0.37 | 0.18–0.74 | 0.005 |
| Amazonian region | 0.26 | 0.11–0.61 | 0.002 |
| **Macroregion (ref: Metropolitan Lima)** |  |  |  |
| North | 0.35 | 0.17–0.73 | 0.005 |
| Central | 0.27 | 0.13–0.56 | <0.001 |
| South | 0.58 | 0.29–1.18 | 0.132 |
| Oriente | 0.28 | 0.10–0.76 | 0.013 |

**Panel B. Annual psychology service use**

| **Variable** | **cPR** | **95% CI** | **p** |
| --- | --- | --- | --- |
| Age (continuous) | 0.98 | 0.97–0.99 | <0.001 |
| **Sex** |  |  |  |
| Female vs male | 2.03 | 1.47–2.80 | <0.001 |
| **Educational attainment (ref: complete secondary)** |  |  |  |
| No education | 0.03 | 0.01–0.18 | <0.001 |
| Less than complete secondary | 0.62 | 0.45–0.84 | 0.002 |
| Higher education | 1.32 | 0.97–1.79 | 0.081 |
| **Marital status (ref: single)** |  |  |  |
| Married | 0.61 | 0.42–0.87 | 0.006 |
| Separated | 1.14 | 0.72–1.79 | 0.576 |
| Divorced | 1.73 | 0.68–4.36 | 0.247 |
| Widowed | 0.75 | 0.32–1.74 | 0.503 |
| Cohabiting | 0.65 | 0.47–0.90 | 0.009 |
| **Poverty status (ref: not poor)** |  |  |  |
| Poor | 0.62 | 0.47–0.81 | <0.001 |
| Extreme poverty | 0.26 | 0.16–0.45 | <0.001 |
| **Area of residence (ref: urban)** |  |  |  |
| Rural | 0.36 | 0.24–0.55 | <0.001 |
| **Natural region (ref: Metropolitan Lima)** |  |  |  |
| Coast | 1.40 | 0.95–2.05 | 0.087 |
| Highlands | 0.98 | 0.66–1.47 | 0.928 |
| Amazonian region | 0.70 | 0.44–1.11 | 0.127 |
| **Macroregion (ref: Metropolitan Lima)** |  |  |  |
| North | 1.07 | 0.69–1.67 | 0.749 |
| Central | 1.16 | 0.78–1.72 | 0.456 |
| South | 1.28 | 0.84–1.93 | 0.249 |
| Oriente | 0.63 | 0.38–1.06 | 0.080 |

**Panel C. High satisfaction with psychiatry care**

| **Variable** | **cPR** | **95% CI** | **p** |
| --- | --- | --- | --- |
| Age (continuous) | 1.01 | 1.00–1.03 | 0.138 |
| **Sex** |  |  |  |
| Female vs male | 1.24 | 0.79–1.95 | 0.350 |
| **Educational attainment (ref: complete secondary)** |  |  |  |
| No education | — | — | — |
| Less than complete secondary | 0.73 | 0.33–1.65 | 0.455 |
| Higher education | 1.44 | 0.96–2.14 | 0.075 |
| **Marital status (ref: single)** |  |  |  |
| Married | 1.33 | 0.89–2.01 | 0.165 |
| Separated | 0.46 | 0.14–1.51 | 0.200 |
| Divorced | 1.57 | 1.09–2.25 | 0.015 |
| Widowed | 1.54 | 1.07–2.22 | 0.021 |
| Cohabiting | 1.27 | 0.79–2.02 | 0.319 |
| **Poverty status (ref: not poor)** |  |  |  |
| Poor | 1.08 | 0.76–1.53 | 0.683 |
| Extreme poverty | 1.36 | 1.05–1.76 | 0.021 |
| **Area of residence (ref: urban)** |  |  |  |
| Rural | 1.11 | 0.69–1.80 | 0.660 |
| **Natural region (ref: Metropolitan Lima)** |  |  |  |
| Coast | 1.00 | 0.67–1.52 | 0.982 |
| Highlands | 0.85 | 0.51–1.41 | 0.518 |
| Amazonian region | 1.22 | 0.83–1.80 | 0.304 |
| **Macroregion (ref: Metropolitan Lima)** |  |  |  |
| North | 0.84 | 0.50–1.41 | 0.501 |
| Central | 1.13 | 0.76–1.69 | 0.554 |
| South | 0.91 | 0.55–1.49 | 0.701 |
| Oriente | 1.29 | 0.89–1.86 | 0.176 |

**Panel D. High satisfaction with psychology care**

| **Variable** | **cPR** | **95% CI** | **p** |
| --- | --- | --- | --- |
| Age (continuous) | 1.01 | 1.00–1.01 | 0.083 |
| **Sex** |  |  |  |
| Female vs male | 1.01 | 0.84–1.21 | 0.939 |
| **Educational attainment (ref: complete secondary)** |  |  |  |
| No education | 1.36 | 1.20–1.54 | <0.001 |
| Less than complete secondary | 1.05 | 0.87–1.26 | 0.603 |
| Higher education | 1.08 | 0.90–1.31 | 0.407 |
| **Marital status (ref: single)** |  |  |  |
| Married | 1.03 | 0.83–1.27 | 0.775 |
| Separated | 1.07 | 0.78–1.47 | 0.686 |
| Divorced | 0.78 | 0.33–1.81 | 0.561 |
| Widowed | 0.98 | 0.54–1.77 | 0.948 |
| Cohabiting | 1.06 | 0.88–1.28 | 0.535 |
| **Poverty status (ref: not poor)** |  |  |  |
| Poor | 1.15 | 1.01–1.31 | 0.035 |
| Extreme poverty | 1.03 | 0.76–1.41 | 0.842 |
| **Area of residence (ref: urban)** |  |  |  |
| Rural | 1.19 | 1.04–1.35 | 0.011 |
| **Natural region (ref: Metropolitan Lima)** |  |  |  |
| Coast | 1.20 | 0.92–1.57 | 0.170 |
| Highlands | 1.19 | 0.91–1.55 | 0.212 |
| Amazonian region | 1.22 | 0.93–1.61 | 0.158 |
| **Macroregion (ref: Metropolitan Lima)** |  |  |  |
| North | 1.22 | 0.92–1.60 | 0.164 |
| Central | 1.22 | 0.93–1.60 | 0.144 |
| South | 1.12 | 0.84–1.48 | 0.441 |
| Oriente | 1.37 | 1.05–1.78 | 0.018 |

**Panel E. Psychiatry care perceived as very helpful**

| **Variable** | **cPR** | **95% CI** | **p** |
| --- | --- | --- | --- |
| Age (continuous) | 1.01 | 0.99–1.04 | 0.318 |
| **Sex** |  |  |  |
| Female vs male | 1.36 | 0.66–2.77 | 0.404 |
| **Educational attainment (ref: complete secondary)** |  |  |  |
| No education | — | — | — |
| Less than complete secondary | 0.45 | 0.14–1.42 | 0.175 |
| Higher education | 2.02 | 1.15–3.53 | 0.014 |
| **Marital status (ref: single)** |  |  |  |
| Married | 1.62 | 0.89–2.95 | 0.116 |
| Separated | 0.50 | 0.13–1.88 | 0.306 |
| Divorced | 1.50 | 0.55–4.11 | 0.432 |
| Widowed | 2.27 | 1.41–3.65 | 0.001 |
| Cohabiting | 0.89 | 0.34–2.33 | 0.818 |
| **Poverty status (ref: not poor)** |  |  |  |
| Poor | 0.75 | 0.36–1.57 | 0.441 |
| Extreme poverty | 1.77 | 1.25–2.51 | 0.001 |
| **Area of residence (ref: urban)** |  |  |  |
| Rural | 1.08 | 0.45–2.56 | 0.864 |
| **Natural region (ref: Metropolitan Lima)** |  |  |  |
| Coast | 0.84 | 0.46–1.52 | 0.555 |
| Highlands | 0.55 | 0.26–1.16 | 0.117 |
| Amazonian region | 1.34 | 0.76–2.34 | 0.312 |
| **Macroregion (ref: Metropolitan Lima)** |  |  |  |
| North | 0.77 | 0.37–1.59 | 0.480 |
| Central | 0.95 | 0.51–1.78 | 0.881 |
| South | 0.51 | 0.23–1.10 | 0.086 |
| Oriente | 1.62 | 1.00–2.61 | 0.048 |

**Panel F. Psychology care perceived as very helpful**

| **Variable** | **cPR** | **95% CI** | **p** |
| --- | --- | --- | --- |
| Age (continuous) | 1.00 | 0.99–1.01 | 0.756 |
| **Sex** |  |  |  |
| Female vs male | 1.10 | 0.80–1.52 | 0.555 |
| **Educational attainment (ref: complete secondary)** |  |  |  |
| No education | 2.03 | 1.67–2.46 | <0.001 |
| Less than complete secondary | 1.05 | 0.77–1.42 | 0.777 |
| Higher education | 1.14 | 0.85–1.52 | 0.394 |
| **Marital status (ref: single)** |  |  |  |
| Married | 0.96 | 0.68–1.34 | 0.788 |
| Separated | 0.92 | 0.59–1.43 | 0.703 |
| Divorced | 1.06 | 0.45–2.51 | 0.893 |
| Widowed | 0.97 | 0.43–2.18 | 0.944 |
| Cohabiting | 0.87 | 0.63–1.20 | 0.398 |
| **Poverty status (ref: not poor)** |  |  |  |
| Poor | 1.14 | 0.88–1.46 | 0.316 |
| Extreme poverty | 0.84 | 0.48–1.48 | 0.552 |
| **Area of residence (ref: urban)** |  |  |  |
| Rural | 1.38 | 1.06–1.79 | 0.015 |
| **Natural region (ref: Metropolitan Lima)** |  |  |  |
| Coast | 1.54 | 1.01–2.35 | 0.046 |
| Highlands | 1.20 | 0.76–1.89 | 0.431 |
| Amazonian region | 1.44 | 0.90–2.30 | 0.132 |
| **Macroregion (ref: Metropolitan Lima)** |  |  |  |
| North | 1.56 | 1.00–2.43 | 0.050 |
| Central | 1.45 | 0.94–2.26 | 0.095 |
| South | 1.15 | 0.72–1.84 | 0.559 |
| Oriente | 1.54 | 0.95–2.49 | 0.081 |

**Panel G. Lifetime mental health telephone-line use**

| **Variable** | **cPR** | **95% CI** | **p** |
| --- | --- | --- | --- |
| Age (continuous) | 0.99 | 0.95–1.03 | 0.624 |
| **Sex** |  |  |  |
| Female vs male | 1.61 | 0.63–4.10 | 0.319 |
| **Educational attainment (ref: complete secondary)** |  |  |  |
| No education | — | — | <0.001* |
| Less than complete secondary | 0.57 | 0.24–1.35 | 0.199 |
| Higher education | 1.42 | 0.53–3.78 | 0.484 |
| **Marital status (ref: single)** |  |  |  |
| Married | 0.44 | 0.13–1.51 | 0.191 |
| Separated | 1.18 | 0.43–3.26 | 0.754 |
| Divorced | 1.64 | 0.20–13.55 | 0.646 |
| Widowed | 2.40 | 0.42–13.62 | 0.324 |
| Cohabiting | 0.36 | 0.14–0.94 | 0.038 |
| **Poverty status (ref: not poor)** |  |  |  |
| Poor | 0.67 | 0.26–1.73 | 0.410 |
| Extreme poverty | 0.37 | 0.12–1.13 | 0.080 |
| **Area of residence (ref: urban)** |  |  |  |
| Rural | 0.20 | 0.07–0.57 | 0.003 |
| **Natural region (ref: Metropolitan Lima)** |  |  |  |
| Coast | 0.21 | 0.08–0.57 | 0.002 |
| Highlands | 0.38 | 0.15–0.94 | 0.037 |
| Amazonian region | 0.23 | 0.09–0.61 | 0.003 |
| **Macroregion (ref: Metropolitan Lima)** |  |  |  |
| North | 0.14 | 0.03–0.58 | 0.007 |
| Central | 0.25 | 0.09–0.73 | 0.011 |
| South | 0.51 | 0.20–1.26 | 0.143 |
| Oriente | 0.30 | 0.11–0.84 | 0.022 |

cPR, crude prevalence ratio; CI, confidence interval. Poisson regression models with a log link and linearized standard errors accounted for the complex ENSM Peru 2022 design. p values were based on design-adjusted t statistics; reference categories are shown in parentheses; NE denotes not estimable. High satisfaction combined satisfied and very satisfied responses and was compared with all lower categories. Care perceived as very helpful was compared with not at all, a little, or moderately helpful. Satisfaction and perceived-helpfulness models were estimated as subpopulation analyses among users of the corresponding service during the previous 12 months. Telephone-line use denotes lifetime use and is not restricted to the previous 12 months. NE cells reflect absent events, insufficient observations, or model nonconvergence. Psychiatry experience estimates were based on 120 users and should be interpreted cautiously. ENSM, National Mental Health Survey.

**Supplementary Table S3. Exploratory indicators of clinical complexity and annual contact gaps among adults meeting survey-based criteria for at least one 12-month mental disorder, ENSM Peru 2022**

*Weighted secondary analyses; these indicators are not measures of severity, persistence, or comprehensive clinical need*

**Panel A. Recent diagnostic activity**

| **Category** | **Total n** | **n with evaluable contact gap** | **Weighted indicator, %** | **95% CI** | **Contact gap, %** | **Contact-gap 95% CI** | **cPR** | **cPR 95% CI** | **p** |
| --- | --- | --- | --- | --- | --- | --- | --- | --- | --- |
| 12-month disorder without 30-day disorder | 1077 | 1038 | 64.3 | 60.2–68.2 | 87.0 | 83.6–90.4 | Reference | — | — |
| 12-month and 30-day disorder | 631 | 611 | 35.7 | 31.8–39.8 | 80.0 | 74.0–85.9 | 0.92 | 0.85–1.00 | 0.047 |

**Panel B. Partial comorbidity across five available diagnostic domains**

| **Category** | **Total n** | **n with evaluable contact gap** | **Weighted indicator, %** | **95% CI** | **Contact gap, %** | **Contact-gap 95% CI** | **cPR** | **cPR 95% CI** | **p** |
| --- | --- | --- | --- | --- | --- | --- | --- | --- | --- |
| One positive domain | 1127 | 1122 | 82.9 | 79.6–85.7 | 87.3 | 84.6–90.1 | Reference | — | — |
| Two or more positive domains | 240 | 239 | 17.1 | 14.3–20.4 | 71.5 | 62.6–80.4 | 0.82 | 0.72–0.93 | 0.002 |

**Panel C. Recent diagnostic activity by partial comorbidity**

| **Category** | **n** | **Recent diagnostic activity, %** | **95% CI** | **cPR** | **cPR 95% CI** | **p** |
| --- | --- | --- | --- | --- | --- | --- |
| One positive domain | 1127 | 30.1 | 25.6–34.6 | Reference | — | — |
| Two or more positive domains | 240 | 58.9 | 49.9–67.8 | 1.95 | 1.58–2.41 | <0.001 |

Notes. Percentages, confidence intervals, and prevalence ratios account for the complex ENSM Peru 2022 survey design. Poisson models with a log link were crude, weighted, and exploratory.

Recent diagnostic activity indicates that a 12-month disorder was also present during the previous 30 days. It does not measure persistence, chronicity, episode duration, or severity.

The domain measure includes anxiety disorders, affective disorders, psychosis screening, eating disorders, and harmful substance use or dependence. It is not a complete diagnostic count.

The binary domain comparison excluded 198 participants whose 12-month disorder was not represented by the five domains and 143 with incomplete domain information; these groups should not be combined as simple missing data.

The gap is the annual contact gap used in the manuscript. A smaller gap does not establish that contact was adequate, continuous, specific, or effective. The available documentation did not support construction of comprehensive measures of severity, functional impairment, or clinical course. CI, confidence interval; cPR, crude prevalence ratio; ENSM, National Mental Health Survey.

**Supplementary Table S4. Analytic variable dictionary for the ENSM Peru 2022 secondary analysis**

*Source-field names are reproduced exactly as they appeared in the analytic files. Definitions describe the implemented Stata logic. Where questionnaire wording, coding metadata, or a derivation algorithm was unavailable, this is stated explicitly and no equivalence was inferred. This sheet is intentionally unnumbered and may be designated as a supplementary table if requested by the journal.*

| **Analytic construct** | **Original ENSM 2022 variable(s)** | **Analysis variable** | **Operational definition and coding** | **Role in this study** |
| --- | --- | --- | --- | --- |
| **A. Primary 12-month perceived-need measures** |  |  |  |  |
| 12-month mental health-related service contact status | lt_mhs_access_anual | mhs_anual | Supplied binary 12-month contact indicator used for questionnaire routing and contact-gap calculations. Analyses used 1 = contact and 0 = no contact; missing values remained missing. The available documentation did not establish how the supplied indicator was constructed or which sources were included. | Routing variable for the derived perceived-need measures in Tables 4–5; also the broad 12-month contact indicator in Tables 2–3 and Supplementary Table S1. |
| Direct 12-month self-perceived need | demanse | self_need_anual | Direct item asked whether, during the previous 12 months, the respondent felt that they might need to see a professional because of emotional or nervous problems or alcohol/drug use. Valid binary responses were retained when annual contact was absent or contact status was missing. | Source component of need_self_12m; not used alone as the final Tables 4–5 outcome. |
| Direct 12-month need identified by others | demannose | others_need_anual | Direct item (SR124) asked whether, during the previous 12 months, anyone encouraged or pressured the respondent to consult a professional about emotions, mental health, or alcohol/drug use. Valid binary responses were retained when annual contact was absent or contact status was missing. | Source component of need_others_12m; not used alone as the final Tables 4–5 outcome. |
| Reason for seeking care among respondents with annual contact | SR111, supplied as recoded field sr111_r | sr111_r | CAPI routing item for respondents with annual contact. Categories used in the analysis were 1 = respondent wanted to seek care, 2 = someone pressured the respondent, 3 = both, 8 = do not know, and 9 = no response. The literal questionnaire wording was not present in the documentation reviewed. | Source component of both final perceived-need outcomes in Tables 4–5. |
| Encouragement or pressure by others after self-initiated care | SR113, supplied as recoded field sr113_r | sr113_r | CAPI follow-up item used when SR111 = 1. Categories used were 1 = yes, 5 = no, 8 = do not know, and 9 = no response. The literal questionnaire wording was not present in the documentation reviewed. | Distinguishes whether others also encouraged or pressured respondents whose SR111 response indicated self-initiated care; used in need_others_12m. |
| Derived 12-month self-perceived need | lt_mhs_access_anual, demanse, and sr111_r | need_self_12m | Binary final measure. If mhs_anual = 0, or mhs_anual was missing and a valid direct response was available, the value of self_need_anual was retained. If mhs_anual = 1, SR111 values 1 or 3 were coded 1 and value 2 was coded 0. Required do-not-know or nonresponse values remained missing. | Primary self-perceived-need outcome in Tables 4–5. |
| Derived 12-month need identified by others | lt_mhs_access_anual, demannose, sr111_r, and sr113_r | need_others_12m | Binary final measure. If mhs_anual = 0, or mhs_anual was missing and a valid direct response was available, the value of others_need_anual was retained. If mhs_anual = 1, SR111 values 2 or 3 were coded 1; when SR111 = 1, SR113 = 1 was coded 1 and SR113 = 5 was coded 0. Required do-not-know or nonresponse values remained missing. | Primary outcome for need identified by others in Tables 4–5. |
| Derived 12-month any perceived need | need_self_12m and need_others_12m | need_any_12m | Binary derived summary: coded 1 if either final perceived-need measure equaled 1; coded 0 only if both equaled 0; otherwise missing. | Internal descriptive and logical-check variable; not a separately reported final outcome in Tables 4–5. |
| Perceived-need sensitivity variables restricted to known annual-contact status | need_self_12m, need_others_12m, need_any_12m, and mhs_anual | need_self_12m_known; need_others_12m_known; need_any_12m_known | Copies of the corresponding final perceived-need variables, retained only when mhs_anual was observed; set to missing when annual-contact status was missing. | Sensitivity analysis assessing the treatment of missing annual-contact status; documented in the analysis audit rather than the principal tables. |
| **B. Complex survey design variables** |  |  |  |  |
| Final survey weight | raked_weight_trimmed | raked_weight_trimmed | Final probability weight entered as the pweight in all survey analyses. The study used the supplied values and did not assert how the weight was constructed. | Weights all prevalence estimates, confidence intervals, and regression models. |
| Primary sampling unit | ccluster | ccluster | Cluster identifier entered as the first-stage primary sampling unit in the implemented svyset specification. | Accounts for clustering in variance estimation. |
| Dwelling-level sampling unit | dwelling | dwelling | Dwelling identifier entered as the second-stage sampling unit in the implemented svyset specification. | Represents the nested dwelling stage of the complex design. |
| Eligible adults in the dwelling | Num_adults | Num_adults | Source dictionary label: total eligible adults aged 18–59 years in the dwelling. The variable was entered as the third-stage field in the implemented svyset command; no additional interpretation of this dual use was made. | Retained exactly as implemented in the complex survey specification. |
| Survey stratum | Departamento | departamentos_si_callao | Derived 25-category department variable with Lima and Callao kept separate; entered as the stratum variable in svyset. | Defines survey strata for variance estimation. |
| **C. Sociodemographic and geographic variables** |  |  |  |  |
| Age | Age | age | Respondent age in years, analyzed as a continuous variable. | Described in Table 1 and included as a continuous covariate in Tables 3 and 5 and Supplementary Tables S2A–S2B. |
| Detailed educational attainment | Education | education | Supplied categorical educational level. Table 1 reports: no education, preschool, incomplete/complete primary, incomplete/complete secondary, incomplete/complete non-university higher education, incomplete/complete university education, postgraduate education, and bachelor's degree. | Detailed descriptive distribution in Table 1; source for the collapsed model variable edu_cat. |
| Educational attainment used in models | Education | edu_cat | Four-category recode: 0 = no education (source codes 0–1); 1 = less than complete secondary (2–4); 2 = complete secondary, reference category (5, 6, 8, or 11); 3 = higher education (7, 9, or 10). Numeric source codes are reproduced from the implemented do-file. | Covariate in Tables 3 and 5 and Supplementary Tables S2A–S2B. |
| Marital status | civilstatus | eciv | Categorical variable reported as single, married, separated, divorced, widowed, or cohabiting. Single was the reference category in regression models. | Described in Table 1 and used as a covariate in Tables 3 and 5 and Supplementary Tables S2A–S2B. |
| Sex | Genero | sex | Binary sex variable reported as male or female. Male was the reference category in regression models. The available documentation did not support a separate gender-identity measure. | Described in Table 1 and used as a covariate in Tables 3 and 5 and Supplementary Tables S2A–S2B. |
| Poverty status | Pobreza | pobreza | Three-category indicator based on unmet basic needs: not poor, poor, and extreme poverty. Not poor was the reference category in regression models. | Described in Table 1 and used as a covariate in Tables 3 and 5 and Supplementary Tables S2A–S2B. |
| Area of residence | Area | urbano | Binary urban/rural residence indicator. Urban was the reference category in regression models. | Described in Table 1; used for stratified estimates in Tables 2 and 4 and as a covariate in Tables 3 and 5 and Supplementary Tables S2A–S2B. |
| Natural region | Region_4 | regiones | Four-category natural-region variable: Metropolitan Lima, Coast, Highlands, and Amazonian region. Metropolitan Lima was the reference category in regression models. | Described in Table 1; used for stratified estimates in Tables 2 and 4 and as a covariate in Tables 3 and 5 and Supplementary Tables S2A–S2B. |
| Macroregion | Macroregiones | macroregiones | Five-category macroregion variable: Metropolitan Lima, North, Central, South, and Oriente. Metropolitan Lima was the reference category. | Described in Table 1 and used only in crude or sensitivity models in Tables 3 and 5 and Supplementary Tables S2A–S2B. |
| Department, Lima and Callao combined | Departamento_1 | departamentos_no_callao | Derived 24-category department variable with Lima and Callao combined. | Retained for geographic traceability; not used in the final survey-design specification. |
| **D. Survey-based mental-disorder indicators** |  |  |  |  |
| Lifetime any psychiatric disorder | pvtpsq_2 | psiq_vida | Supplied binary or categorical indicator. Analyses treated value 1 as meeting the survey-based definition stated in the construct; the source diagnostic or derivation algorithm was not reconstructed in this project. | Defines the lifetime analytic subpopulation; used in Table 1 and as the denominator for lifetime contact-gap estimates in Tables 2–3 and Supplementary Table S1. |
| 12-month any psychiatric disorder | patpsq_1 | psiq_anual | Supplied binary or categorical indicator. Analyses treated value 1 as meeting the survey-based definition stated in the construct; the source diagnostic or derivation algorithm was not reconstructed in this project. | Defines the 12-month analytic subpopulation; used in Tables 1–5 and Supplementary Tables S1 and S3. |
| 30-day any psychiatric disorder | pmtpsq_1 | psiq_mensual | Supplied binary or categorical indicator. Analyses treated value 1 as meeting the survey-based definition stated in the construct; the source diagnostic or derivation algorithm was not reconstructed in this project. | Used in Table 1 and to derive recent diagnostic activity in Supplementary Table S3. |
| Lifetime anxiety disorder | pvtansiedad | anx_vida | Supplied binary or categorical indicator. Analyses treated value 1 as meeting the survey-based definition stated in the construct; the source diagnostic or derivation algorithm was not reconstructed in this project. | Used for lifetime prevalence in Table 1 and as a disorder-specific denominator for lifetime contact-gap estimates in Table 2. |
| 12-month anxiety disorder | patansiedad | anx_anual | Supplied binary or categorical indicator. Analyses treated value 1 as meeting the survey-based definition stated in the construct; the source diagnostic or derivation algorithm was not reconstructed in this project. | Used for 12-month prevalence in Table 1, as a disorder-specific denominator in Tables 2 and 4, and, where applicable, as an input to Supplementary Table S3. |
| 30-day anxiety disorder | pmtansiedad | anx_mensual | Supplied binary or categorical indicator. Analyses treated value 1 as meeting the survey-based definition stated in the construct; the source diagnostic or derivation algorithm was not reconstructed in this project. | Used for 30-day prevalence in Table 1; retained as supplied for reproducibility. |
| Lifetime affective disorder | pvcta1 | af_vida | Supplied binary or categorical indicator. Analyses treated value 1 as meeting the survey-based definition stated in the construct; the source diagnostic or derivation algorithm was not reconstructed in this project. | Used for lifetime prevalence in Table 1 and as a disorder-specific denominator for lifetime contact-gap estimates in Table 2. |
| 12-month affective disorder | pacta1 | af_anual | Supplied binary or categorical indicator. Analyses treated value 1 as meeting the survey-based definition stated in the construct; the source diagnostic or derivation algorithm was not reconstructed in this project. | Used for 12-month prevalence in Table 1, as a disorder-specific denominator in Tables 2 and 4, and, where applicable, as an input to Supplementary Table S3. |
| 30-day affective disorder | pmcta1 | af_mensual | Supplied binary or categorical indicator. Analyses treated value 1 as meeting the survey-based definition stated in the construct; the source diagnostic or derivation algorithm was not reconstructed in this project. | Used for 30-day prevalence in Table 1; retained as supplied for reproducibility. |
| Lifetime eating disorder | prev_trast_alimentacion_2 | tca_vida | Supplied binary or categorical indicator. Analyses treated value 1 as meeting the survey-based definition stated in the construct; the source diagnostic or derivation algorithm was not reconstructed in this project. | Used for lifetime prevalence in Table 1 and as a disorder-specific denominator for lifetime contact-gap estimates in Table 2. |
| 12-month eating disorder | prev_trast_alimentacion_anual | tca_anual | Supplied binary or categorical indicator. Analyses treated value 1 as meeting the survey-based definition stated in the construct; the source diagnostic or derivation algorithm was not reconstructed in this project. | Used for 12-month prevalence in Table 1, as a disorder-specific denominator in Tables 2 and 4, and, where applicable, as an input to Supplementary Table S3. |
| 30-day eating disorder | prev_trast_alimentacion_mes | tca_mensual | Supplied binary or categorical indicator. Analyses treated value 1 as meeting the survey-based definition stated in the construct; the source diagnostic or derivation algorithm was not reconstructed in this project. | Used for 30-day prevalence in Table 1; retained as supplied for reproducibility. |
| Lifetime psychosis screen | prev_psicosis | psic_vida | Supplied binary screening indicator. Analyses treated value 1 as a positive survey-based psychosis screen for the stated time window; this is not presented as a confirmed psychotic-disorder diagnosis. The source screening algorithm was not reconstructed in this project. | Used for lifetime prevalence in Table 1 and as a disorder-specific denominator for lifetime contact-gap estimates in Table 2. |
| 12-month psychosis screen | prev_anual_psicosis | psic_anual | Supplied binary screening indicator. Analyses treated value 1 as a positive survey-based psychosis screen for the stated time window; this is not presented as a confirmed psychotic-disorder diagnosis. The source screening algorithm was not reconstructed in this project. | Used for 12-month prevalence in Table 1, as a disorder-specific denominator in Tables 2 and 4, and, where applicable, as an input to Supplementary Table S3. |
| Lifetime conduct disorder | Conducta_vida1 | disocial_vida | Supplied binary or categorical indicator. Analyses treated value 1 as meeting the survey-based definition stated in the construct; the source diagnostic or derivation algorithm was not reconstructed in this project. | Retained in the analytic file and documented for reproducibility; not used as a principal variable in the final tables. |
| Lifetime alcohol use | alcoholv | oh_vida | Supplied binary or categorical indicator. Analyses treated value 1 as meeting the survey-based definition stated in the construct; the source diagnostic or derivation algorithm was not reconstructed in this project. | Retained in the analytic file and documented for reproducibility; not used as a principal variable in the final tables. |
| Lifetime alcohol harmful use/dependence | pvundepalcohexC | ohpd_vida | Supplied binary or categorical indicator. Analyses treated value 1 as meeting the survey-based definition stated in the construct; the source diagnostic or derivation algorithm was not reconstructed in this project. | Used for lifetime prevalence in Table 1 and as a disorder-specific denominator for lifetime contact-gap estimates in Table 2. |
| 12-month alcohol harmful use/dependence | paundepalcohexC | ohpd_anual | Supplied binary or categorical indicator. Analyses treated value 1 as meeting the survey-based definition stated in the construct; the source diagnostic or derivation algorithm was not reconstructed in this project. | Used for 12-month prevalence in Table 1, as a disorder-specific denominator in Tables 2 and 4, and, where applicable, as an input to Supplementary Table S3. |
| Lifetime alcohol dependence | pvdepalcohexC | ohd_vida | Supplied binary or categorical indicator. Analyses treated value 1 as meeting the survey-based definition stated in the construct; the source diagnostic or derivation algorithm was not reconstructed in this project. | Retained in the analytic file and documented for reproducibility; not used as a principal variable in the final tables. |
| 12-month alcohol dependence | padepalcohexC | ohd_anual | Supplied binary or categorical indicator. Analyses treated value 1 as meeting the survey-based definition stated in the construct; the source diagnostic or derivation algorithm was not reconstructed in this project. | Retained in the analytic file and documented for reproducibility; not used as a principal variable in the final tables. |
| Lifetime harmful alcohol use | pvunalcohexC | ohp_vida | Supplied binary or categorical indicator. Analyses treated value 1 as meeting the survey-based definition stated in the construct; the source diagnostic or derivation algorithm was not reconstructed in this project. | Retained in the analytic file and documented for reproducibility; not used as a principal variable in the final tables. |
| 12-month harmful alcohol use | paunalcohexC | ohp_anual | Supplied binary or categorical indicator. Analyses treated value 1 as meeting the survey-based definition stated in the construct; the source diagnostic or derivation algorithm was not reconstructed in this project. | Retained in the analytic file and documented for reproducibility; not used as a principal variable in the final tables. |
| Lifetime illegal drug harmful use/dependence | pvundepsiex | ilegal_pod_vida | Supplied binary or categorical indicator. Analyses treated value 1 as meeting the survey-based definition stated in the construct; the source diagnostic or derivation algorithm was not reconstructed in this project. | Used for lifetime prevalence in Table 1 and as a disorder-specific denominator for lifetime contact-gap estimates in Table 2. |
| 12-month illegal drug harmful use/dependence | paundepsiex | ilegal_pod_anual | Supplied binary or categorical indicator. Analyses treated value 1 as meeting the survey-based definition stated in the construct; the source diagnostic or derivation algorithm was not reconstructed in this project. | Used for 12-month prevalence in Table 1, as a disorder-specific denominator in Tables 2 and 4, and, where applicable, as an input to Supplementary Table S3. |
| Lifetime harmful illegal drug use | pvunsipexd | ilegal_cp_vida | Supplied binary or categorical indicator. Analyses treated value 1 as meeting the survey-based definition stated in the construct; the source diagnostic or derivation algorithm was not reconstructed in this project. | Retained in the analytic file and documented for reproducibility; not used as a principal variable in the final tables. |
| 12-month harmful illegal drug use | paunsipexd | ilegal_cp_anual | Supplied binary or categorical indicator. Analyses treated value 1 as meeting the survey-based definition stated in the construct; the source diagnostic or derivation algorithm was not reconstructed in this project. | Retained in the analytic file and documented for reproducibility; not used as a principal variable in the final tables. |
| Lifetime addictive substance harmful use/dependence | pvtundepC | adic_pod_vida | Supplied binary or categorical indicator. Analyses treated value 1 as meeting the survey-based definition stated in the construct; the source diagnostic or derivation algorithm was not reconstructed in this project. | Used for lifetime prevalence in Table 1 and as a disorder-specific denominator for lifetime contact-gap estimates in Table 2. |
| 12-month addictive substance harmful use/dependence | patundepC | adic_pod_anual | Supplied binary or categorical indicator. Analyses treated value 1 as meeting the survey-based definition stated in the construct; the source diagnostic or derivation algorithm was not reconstructed in this project. | Used for 12-month prevalence in Table 1, as a disorder-specific denominator in Tables 2 and 4, and, where applicable, as an input to Supplementary Table S3. |
| Lifetime addictive substance dependence | pvtdepsa | adic_dep_vida | Supplied binary or categorical indicator. Analyses treated value 1 as meeting the survey-based definition stated in the construct; the source diagnostic or derivation algorithm was not reconstructed in this project. | Retained in the analytic file and documented for reproducibility; not used as a principal variable in the final tables. |
| 12-month addictive substance dependence | patdepsa | adic_dep_anual | Supplied binary or categorical indicator. Analyses treated value 1 as meeting the survey-based definition stated in the construct; the source diagnostic or derivation algorithm was not reconstructed in this project. | Retained in the analytic file and documented for reproducibility; not used as a principal variable in the final tables. |
| Lifetime harmful addictive substance use | pvtunsa | adic_cp_vida | Supplied binary or categorical indicator. Analyses treated value 1 as meeting the survey-based definition stated in the construct; the source diagnostic or derivation algorithm was not reconstructed in this project. | Retained in the analytic file and documented for reproducibility; not used as a principal variable in the final tables. |
| 12-month harmful addictive substance use | patunsa | adic_cp_mensual | Supplied binary or categorical indicator. Analyses treated value 1 as meeting the survey-based definition stated in the construct; the source diagnostic or derivation algorithm was not reconstructed in this project. The analysis-variable name contains 'mensual', although the source dictionary and construct describe a 12-month indicator; the name was retained verbatim and should be confirmed before reuse. | Retained for traceability with an explicit time-window/name mismatch; not used as a principal variable in the final tables. |
| **E. Mental health-related contact, provider use, and telephone-line use** |  |  |  |  |
| Lifetime mental health-related service contact | lt_mhs_access | mhs_vida | Supplied binary lifetime contact indicator. Analyses used 1 = contact and 0 = no contact. The available documentation did not establish its full source composition or its equivalence to the provider-specific indicators. | Broad lifetime contact definition in Tables 2–3 and Supplementary Table S1; lifetime estimates are interpreted descriptively. |
| Lifetime use of a mental health telephone line | sr11_r | ever_linea | Direct item asking whether the respondent had ever used a telephone helpline (including Line 113, MINSA, or Noguchi) for emotional or nervous problems. Analyzed as a lifetime binary indicator. | Described in Table 6 and used as an exploratory outcome in Supplementary Table S2B. |
| Timing of most recent mental health telephone-line use | sr11_b | when_linea | Categorical follow-up among lifetime telephone-line users: previous month, 2–6 months ago, 7–12 months ago, more than one year ago, or do not know. | Described among telephone-line users in Table 6. |
| 12-month affective-disorder-specific care | aassmcta | acc_af_anual | Supplied binary indicator of mental health-related care during the previous 12 months for an affective disorder; the source construction algorithm was not re-derived. | Source for the affective-disorder-specific contact-gap sensitivity variable. |
| Lifetime anxiety-disorder-specific care | advssmctans | acc_anx_vida | Supplied binary lifetime indicator of mental health-related care for an anxiety disorder; the source construction algorithm was not re-derived. | Retained for disorder-specific access traceability. |
| 12-month anxiety-disorder-specific care | aassmctans | acc_anx_anual | Supplied binary indicator of mental health-related care during the previous 12 months for an anxiety disorder; the source construction algorithm was not re-derived. | Source for the anxiety-disorder-specific contact-gap sensitivity variable. |
| Lifetime addictive-substance-specific care | avcpdcsa | acc_adic_vida | Supplied binary lifetime indicator of mental health-related care for harmful use of or dependence on any addictive substance; the source construction algorithm was not re-derived. | Retained for disorder-specific access traceability. |
| Lifetime alcohol-specific care | alcoholavtcpdep | acc_ohpd_vida | Supplied binary lifetime indicator of mental health-related care for harmful alcohol use or dependence; the source construction algorithm was not re-derived. | Retained for disorder-specific access traceability. |
| 12-month alcohol-specific care | alcoholaatcp | acc_ohpd_anual | Supplied binary indicator of mental health-related care during the previous 12 months for harmful alcohol use or dependence; the source construction algorithm was not re-derived. | Retained for disorder-specific access traceability. |
| Lifetime eating-disorder-specific care | avssmcondalim_C | acc_tca_vida | Supplied binary lifetime indicator of mental health-related care for an eating disorder; the source construction algorithm was not re-derived. | Retained for disorder-specific access traceability. |
| Lifetime psychiatry service use, population | lt_psychiat_acc_p | acc_psiq_pob_vida | Supplied binary population-level indicator of lifetime psychiatry service use. | Provider component of Supplementary Table S1 contact definitions. |
| Lifetime psychiatry service use, users | lt_psychiat_acc_u | acc_psiq_usu_vida | Supplied lifetime psychiatry-use indicator defined in the source file among service users; the exact denominator was not reconstructed. | Retained for provider-use traceability; not used as a principal final-table variable. |
| 12-month psychiatry service use, population | py_psychiat_acc_p | acc_psq_pob_anual | Supplied binary population-level indicator of psychiatry service use during the previous 12 months. | Described in Table 6; outcome in Supplementary Tables S2A–S2B and provider component of Supplementary Table S1. |
| Lifetime psychology service use, population | lt_psycho_acc_p | acc_psc_pob_vida | Supplied binary population-level indicator of lifetime psychology service use. | Provider component of Supplementary Table S1 contact definitions. |
| Lifetime psychology service use, users | lt_psycho_acc_u | acc_psc_usu_vida | Supplied lifetime psychology-use indicator defined in the source file among service users; the exact denominator was not reconstructed. | Retained for provider-use traceability; not used as a principal final-table variable. |
| 12-month psychology service use, population | py_psycho_acc_p | acc_psc_pob_anual | Supplied binary population-level indicator of psychology service use during the previous 12 months. | Described in Table 6; outcome in Supplementary Tables S2A–S2B and provider component of Supplementary Table S1. |
| Lifetime general-practitioner or other medical-specialist contact | lt_gpra_ospec_acc_p | lt_gpra_ospec_acc_p | Supplied binary population-level provider-contact indicator. Literal questionnaire wording and derivation metadata were not available in the documentation reviewed. | Provider component of the specialist-plus-physician and expanded health-sector definitions in Supplementary Table S1. |
| 12-month general-practitioner or other medical-specialist contact | py_gpra_ospec_acc_p | py_gpra_ospec_acc_p | Supplied binary population-level provider-contact indicator for the previous 12 months. Literal questionnaire wording and derivation metadata were not available in the documentation reviewed. | Provider component of the specialist-plus-physician and expanded health-sector definitions in Supplementary Table S1. |
| Lifetime contact with another health professional | lt_otherhp_acc_p | lt_otherhp_acc_p | Supplied binary population-level provider-contact indicator. Literal questionnaire wording and derivation metadata were not available in the documentation reviewed. | Additional provider component of the expanded health-sector definition in Supplementary Table S1. |
| 12-month contact with another health professional | py_otherhp_acc_p | py_otherhp_acc_p | Supplied binary population-level provider-contact indicator for the previous 12 months. Literal questionnaire wording and derivation metadata were not available in the documentation reviewed. | Additional provider component of the expanded health-sector definition in Supplementary Table S1. |
| Lifetime specialist contact | acc_psiq_pob_vida and acc_psc_pob_vida | contacto_esp_vida | Row maximum of the lifetime psychiatry and psychology indicators. Coded 1 when either component equaled 1, 0 when all observed components equaled 0, and missing only when all components were missing. | Alternative lifetime contact definition in Supplementary Table S1. |
| 12-month specialist contact | acc_psq_pob_anual and acc_psc_pob_anual | contacto_esp_anual | Row maximum of the 12-month psychiatry and psychology indicators. Coded 1 when either component equaled 1, 0 when all observed components equaled 0, and missing only when all components were missing. | Alternative 12-month contact definition in Supplementary Table S1. |
| Lifetime specialist-plus-physician contact | acc_psiq_pob_vida, acc_psc_pob_vida, and lt_gpra_ospec_acc_p | contacto_salud_core_vida | Row maximum of lifetime psychiatry, psychology, and general-practitioner/other-medical-specialist indicators. | Alternative lifetime contact definition in Supplementary Table S1. |
| 12-month specialist-plus-physician contact | acc_psq_pob_anual, acc_psc_pob_anual, and py_gpra_ospec_acc_p | contacto_salud_core_anual | Row maximum of 12-month psychiatry, psychology, and general-practitioner/other-medical-specialist indicators. | Alternative 12-month contact definition in Supplementary Table S1. |
| Lifetime expanded health-sector contact | acc_psiq_pob_vida, acc_psc_pob_vida, lt_gpra_ospec_acc_p, and lt_otherhp_acc_p | contacto_salud_ext_vida | Row maximum of the four lifetime health-provider indicators. | Alternative lifetime contact definition in Supplementary Table S1. |
| 12-month expanded health-sector contact | acc_psq_pob_anual, acc_psc_pob_anual, py_gpra_ospec_acc_p, and py_otherhp_acc_p | contacto_salud_ext_anual | Row maximum of the four 12-month health-provider indicators. | Alternative 12-month contact definition in Supplementary Table S1. |
| Conservative lifetime expanded health-sector contact | contacto_salud_ext_vida plus component-missingness count | contacto_salud_ext_vida_cons | Copy of contacto_salud_ext_vida, except apparent zeros were set to missing if one or more component provider indicators were missing. | Missing-data sensitivity analysis for Supplementary Table S1. |
| Conservative 12-month expanded health-sector contact | contacto_salud_ext_anual plus component-missingness count | contacto_salud_ext_anual_cons | Copy of contacto_salud_ext_anual, except apparent zeros were set to missing if one or more component provider indicators were missing. | Missing-data sensitivity analysis for Supplementary Table S1; the 12-month result is summarized in the table note. |
| **F. Contact-gap outcomes** |  |  |  |  |
| Lifetime contact gap, any mental disorder | lt_mhs_access and pvtpsq_2 | bch_psiq_vida | Binary derived outcome defined only among participants positive for any lifetime mental disorder (psiq_vida = 1). Coded 1 when mhs_vida = 0 (no contact) and 0 when mhs_vida = 1 (contact); otherwise missing. This is a contact-gap indicator and does not measure adequacy, continuity, specificity, or effectiveness of care. | Principal lifetime contact-gap outcome in Tables 2–3. |
| 12-month contact gap, any mental disorder | lt_mhs_access_anual and patpsq_1 | bch_psiq_anual | Binary derived outcome defined only among participants positive for any 12-month mental disorder (psiq_anual = 1). Coded 1 when mhs_anual = 0 (no contact) and 0 when mhs_anual = 1 (contact); otherwise missing. This is a contact-gap indicator and does not measure adequacy, continuity, specificity, or effectiveness of care. | Principal 12-month contact-gap outcome in Tables 2–3 and Supplementary Table S3. |
| 12-month contact gap, affective disorders | pacta1 and lt_mhs_access_anual | bch_af_anual | Binary derived outcome defined only among participants positive for a 12-month affective disorder (af_anual = 1). Coded 1 when mhs_anual = 0 (no contact) and 0 when mhs_anual = 1 (contact); otherwise missing. This is a contact-gap indicator and does not measure adequacy, continuity, specificity, or effectiveness of care. | Disorder-specific outcome in Table 2. |
| 12-month specific-care gap, affective disorders | pacta1 and aassmcta | bch_af_anual_esp | Binary derived outcome defined only among participants positive for a 12-month affective disorder (af_anual = 1). Coded 1 when acc_af_anual = 0 (no contact) and 0 when acc_af_anual = 1 (contact); otherwise missing. This is a contact-gap indicator and does not measure adequacy, continuity, specificity, or effectiveness of care. | Sensitivity variable retained for disorder-specific access traceability. |
| Lifetime contact gap, affective disorders | pvcta1 and lt_mhs_access | bch_af_vida | Binary derived outcome defined only among participants positive for a lifetime affective disorder (af_vida = 1). Coded 1 when mhs_vida = 0 (no contact) and 0 when mhs_vida = 1 (contact); otherwise missing. This is a contact-gap indicator and does not measure adequacy, continuity, specificity, or effectiveness of care. | Disorder-specific outcome in Table 2. |
| 12-month contact gap, anxiety disorders | patansiedad and lt_mhs_access_anual | bch_anx_anual | Binary derived outcome defined only among participants positive for a 12-month anxiety disorder (anx_anual = 1). Coded 1 when mhs_anual = 0 (no contact) and 0 when mhs_anual = 1 (contact); otherwise missing. This is a contact-gap indicator and does not measure adequacy, continuity, specificity, or effectiveness of care. | Disorder-specific outcome in Table 2. |
| 12-month specific-care gap, anxiety disorders | patansiedad and aassmctans | bch_anx_anual_esp | Binary derived outcome defined only among participants positive for a 12-month anxiety disorder (anx_anual = 1). Coded 1 when acc_anx_anual = 0 (no contact) and 0 when acc_anx_anual = 1 (contact); otherwise missing. This is a contact-gap indicator and does not measure adequacy, continuity, specificity, or effectiveness of care. | Sensitivity variable retained for disorder-specific access traceability. |
| Lifetime contact gap, anxiety disorders | pvtansiedad and lt_mhs_access | bch_anx_vida | Binary derived outcome defined only among participants positive for a lifetime anxiety disorder (anx_vida = 1). Coded 1 when mhs_vida = 0 (no contact) and 0 when mhs_vida = 1 (contact); otherwise missing. This is a contact-gap indicator and does not measure adequacy, continuity, specificity, or effectiveness of care. | Disorder-specific outcome in Table 2. |
| 12-month contact gap, eating disorders | prev_trast_alimentacion_anual and lt_mhs_access_anual | bch_tca_anual | Binary derived outcome defined only among participants positive for a 12-month eating disorder (tca_anual = 1). Coded 1 when mhs_anual = 0 (no contact) and 0 when mhs_anual = 1 (contact); otherwise missing. This is a contact-gap indicator and does not measure adequacy, continuity, specificity, or effectiveness of care. | Disorder-specific outcome in Table 2. |
| Lifetime contact gap, eating disorders | prev_trast_alimentacion_2 and lt_mhs_access | bch_tca_vida | Binary derived outcome defined only among participants positive for a lifetime eating disorder (tca_vida = 1). Coded 1 when mhs_vida = 0 (no contact) and 0 when mhs_vida = 1 (contact); otherwise missing. This is a contact-gap indicator and does not measure adequacy, continuity, specificity, or effectiveness of care. | Disorder-specific outcome in Table 2. |
| 12-month contact gap, positive psychosis screen | prev_anual_psicosis and lt_mhs_access_anual | bch_psic_anual | Binary derived outcome defined only among participants positive for a positive 12-month psychosis screen (psic_anual = 1). Coded 1 when mhs_anual = 0 (no contact) and 0 when mhs_anual = 1 (contact); otherwise missing. This is a contact-gap indicator and does not measure adequacy, continuity, specificity, or effectiveness of care. | Screen-specific outcome in Table 2; not interpreted as a confirmed psychotic-disorder treatment gap. |
| Lifetime contact gap, positive psychosis screen | prev_psicosis and lt_mhs_access | bch_psic_vida | Binary derived outcome defined only among participants positive for a positive lifetime psychosis screen (psic_vida = 1). Coded 1 when mhs_vida = 0 (no contact) and 0 when mhs_vida = 1 (contact); otherwise missing. This is a contact-gap indicator and does not measure adequacy, continuity, specificity, or effectiveness of care. | Screen-specific outcome in Table 2; not interpreted as a confirmed psychotic-disorder treatment gap. |
| 12-month contact gap, harmful alcohol use or dependence | paundepalcohexC and lt_mhs_access_anual | bch_ohpd_anual | Binary derived outcome defined only among participants positive for 12-month harmful alcohol use or dependence (ohpd_anual = 1). Coded 1 when mhs_anual = 0 (no contact) and 0 when mhs_anual = 1 (contact); otherwise missing. This is a contact-gap indicator and does not measure adequacy, continuity, specificity, or effectiveness of care. | Disorder-specific outcome in Table 2. |
| Lifetime contact gap, harmful alcohol use or dependence | pvundepalcohexC and lt_mhs_access | bch_ohpd_vida | Binary derived outcome defined only among participants positive for lifetime harmful alcohol use or dependence (ohpd_vida = 1). Coded 1 when mhs_vida = 0 (no contact) and 0 when mhs_vida = 1 (contact); otherwise missing. This is a contact-gap indicator and does not measure adequacy, continuity, specificity, or effectiveness of care. | Disorder-specific outcome in Table 2. |
| Lifetime contact gap, harmful addictive-substance use or dependence | pvtundepC and lt_mhs_access | bch_adic_pod_vida | Binary derived outcome defined only among participants positive for lifetime harmful use of or dependence on an addictive substance (adic_pod_vida = 1). Coded 1 when mhs_vida = 0 (no contact) and 0 when mhs_vida = 1 (contact); otherwise missing. This is a contact-gap indicator and does not measure adequacy, continuity, specificity, or effectiveness of care. | Disorder-specific outcome in Table 2. |
| 12-month contact gap, harmful addictive-substance use or dependence | patundepC and lt_mhs_access_anual | bch_adic_pod_anual | Binary derived outcome defined only among participants positive for 12-month harmful use of or dependence on an addictive substance (adic_pod_anual = 1). Coded 1 when mhs_anual = 0 (no contact) and 0 when mhs_anual = 1 (contact); otherwise missing. This is a contact-gap indicator and does not measure adequacy, continuity, specificity, or effectiveness of care. | Disorder-specific outcome in Table 2. |
| Lifetime contact gap, harmful illegal-substance use or dependence | pvundepsiex and lt_mhs_access | bch_ilegal_pod_vida | Binary derived outcome defined only among participants positive for lifetime harmful use of or dependence on an illegal substance (ilegal_pod_vida = 1). Coded 1 when mhs_vida = 0 (no contact) and 0 when mhs_vida = 1 (contact); otherwise missing. This is a contact-gap indicator and does not measure adequacy, continuity, specificity, or effectiveness of care. | Disorder-specific outcome in Table 2. |
| 12-month contact gap, harmful illegal-substance use or dependence | paundepsiex and lt_mhs_access_anual | bch_ilegal_pod_anual | Binary derived outcome defined only among participants positive for 12-month harmful use of or dependence on an illegal substance (ilegal_pod_anual = 1). Coded 1 when mhs_anual = 0 (no contact) and 0 when mhs_anual = 1 (contact); otherwise missing. This is a contact-gap indicator and does not measure adequacy, continuity, specificity, or effectiveness of care. | Disorder-specific outcome in Table 2. |
| Lifetime broad contact gap | mhs_vida | brecha_amplia_vida | Binary complement of mhs_vida: coded 1 when mhs_vida = 0 (no contact), 0 when mhs_vida = 1 (contact), and missing otherwise. Supplementary Table S1 estimated this lifetime broad-contact outcome within the relevant disorder-defined subpopulation. It does not measure adequacy, continuity, specificity, or effectiveness of care. | Broad lifetime definition in Supplementary Table S1. |
| 12-month broad contact gap | mhs_anual | brecha_amplia_anual | Binary complement of mhs_anual: coded 1 when mhs_anual = 0 (no contact), 0 when mhs_anual = 1 (contact), and missing otherwise. Supplementary Table S1 estimated this 12-month broad-contact outcome within the relevant disorder-defined subpopulation. It does not measure adequacy, continuity, specificity, or effectiveness of care. | Broad 12-month definition in Supplementary Table S1. |
| Lifetime specialist contact gap | contacto_esp_vida | brecha_esp_vida | Binary complement of contacto_esp_vida: coded 1 when contacto_esp_vida = 0 (no contact), 0 when contacto_esp_vida = 1 (contact), and missing otherwise. Supplementary Table S1 estimated this lifetime specialist-contact outcome within the relevant disorder-defined subpopulation. It does not measure adequacy, continuity, specificity, or effectiveness of care. | Alternative lifetime definition in Supplementary Table S1. |
| 12-month specialist contact gap | contacto_esp_anual | brecha_esp_anual | Binary complement of contacto_esp_anual: coded 1 when contacto_esp_anual = 0 (no contact), 0 when contacto_esp_anual = 1 (contact), and missing otherwise. Supplementary Table S1 estimated this 12-month specialist-contact outcome within the relevant disorder-defined subpopulation. It does not measure adequacy, continuity, specificity, or effectiveness of care. | Alternative 12-month definition in Supplementary Table S1. |
| Lifetime specialist-plus-physician contact gap | contacto_salud_core_vida | brecha_salud_core_vida | Binary complement of contacto_salud_core_vida: coded 1 when contacto_salud_core_vida = 0 (no contact), 0 when contacto_salud_core_vida = 1 (contact), and missing otherwise. Supplementary Table S1 estimated this lifetime specialist-plus-physician-contact outcome within the relevant disorder-defined subpopulation. It does not measure adequacy, continuity, specificity, or effectiveness of care. | Alternative lifetime definition in Supplementary Table S1. |
| 12-month specialist-plus-physician contact gap | contacto_salud_core_anual | brecha_salud_core_anual | Binary complement of contacto_salud_core_anual: coded 1 when contacto_salud_core_anual = 0 (no contact), 0 when contacto_salud_core_anual = 1 (contact), and missing otherwise. Supplementary Table S1 estimated this 12-month specialist-plus-physician-contact outcome within the relevant disorder-defined subpopulation. It does not measure adequacy, continuity, specificity, or effectiveness of care. | Alternative 12-month definition in Supplementary Table S1. |
| Lifetime expanded health-sector contact gap | contacto_salud_ext_vida | brecha_salud_ext_vida | Binary complement of contacto_salud_ext_vida: coded 1 when contacto_salud_ext_vida = 0 (no contact), 0 when contacto_salud_ext_vida = 1 (contact), and missing otherwise. Supplementary Table S1 estimated this lifetime expanded-health-sector-contact outcome within the relevant disorder-defined subpopulation. It does not measure adequacy, continuity, specificity, or effectiveness of care. | Alternative lifetime definition in Supplementary Table S1. |
| 12-month expanded health-sector contact gap | contacto_salud_ext_anual | brecha_salud_ext_anual | Binary complement of contacto_salud_ext_anual: coded 1 when contacto_salud_ext_anual = 0 (no contact), 0 when contacto_salud_ext_anual = 1 (contact), and missing otherwise. Supplementary Table S1 estimated this 12-month expanded-health-sector-contact outcome within the relevant disorder-defined subpopulation. It does not measure adequacy, continuity, specificity, or effectiveness of care. | Alternative 12-month definition in Supplementary Table S1. |
| Conservative lifetime expanded health-sector contact gap | contacto_salud_ext_vida_cons | brecha_salud_ext_vida_cons | Binary complement of contacto_salud_ext_vida_cons: coded 1 when contacto_salud_ext_vida_cons = 0 (no contact), 0 when contacto_salud_ext_vida_cons = 1 (contact), and missing otherwise. Supplementary Table S1 estimated this conservative lifetime expanded-health-sector-contact outcome within the relevant disorder-defined subpopulation. It does not measure adequacy, continuity, specificity, or effectiveness of care. | Missing-data sensitivity analysis for Supplementary Table S1. |
| Conservative 12-month expanded health-sector contact gap | contacto_salud_ext_anual_cons | brecha_salud_ext_anual_cons | Binary complement of contacto_salud_ext_anual_cons: coded 1 when contacto_salud_ext_anual_cons = 0 (no contact), 0 when contacto_salud_ext_anual_cons = 1 (contact), and missing otherwise. Supplementary Table S1 estimated this conservative 12-month expanded-health-sector-contact outcome within the relevant disorder-defined subpopulation. It does not measure adequacy, continuity, specificity, or effectiveness of care. | Missing-data sensitivity analysis summarized in Supplementary Table S1. |
| **G. Service experience measures** |  |  |  |  |
| Satisfaction with psychiatry care during the previous 12 months | percpsychipyr | satisf_psiq_anual | Supplied five-level response among psychiatry users: very dissatisfied, dissatisfied, neither satisfied nor dissatisfied, satisfied, or very satisfied. | Described among users in Table 6; source for the dichotomous satisfaction outcome in Supplementary Table S2B. |
| Psychiatry care perceived as helpful during the previous 12 months | perpsychihelppyr | percep_psiq_anual / percep_psq_anual | Supplied ordered response among psychiatry users: not at all, a little, moderately, or a lot. The analysis files contain an unresolved naming inconsistency between percep_psiq_anual and percep_psq_anual; this is reported rather than silently harmonized. | Described in Table 6 and dichotomized for Supplementary Table S2B; confirm the analytic alias before rerunning the model. |
| Satisfaction with psychology care during the previous 12 months | percpsychopyr | satisf_psc_anual | Supplied five-level response among psychology users: very dissatisfied, dissatisfied, neither satisfied nor dissatisfied, satisfied, or very satisfied. | Described among users in Table 6; source for the dichotomous satisfaction outcome in Supplementary Table S2B. |
| Psychology care perceived as helpful during the previous 12 months | perpsychohelppyr | percep_psc_anual | Supplied ordered response among psychology users: not at all, a little, moderately, or a lot. | Described in Table 6 and dichotomized for Supplementary Table S2B. |
| High satisfaction with psychiatry care | percpsychipyr | satisf_psq_anual_cat | Binary recode of satisf_psiq_anual: source categories 0–2 = 0 (very dissatisfied, dissatisfied, or neutral) and categories 3–4 = 1 (satisfied or very satisfied). | Exploratory outcome among 12-month psychiatry users in Supplementary Table S2B. |
| High satisfaction with psychology care | percpsychopyr | satisf_psc_anual_cat | Binary recode of satisf_psc_anual: source categories 0–2 = 0 (very dissatisfied, dissatisfied, or neutral) and categories 3–4 = 1 (satisfied or very satisfied). | Exploratory outcome among 12-month psychology users in Supplementary Table S2B. |
| Psychiatry care perceived as very helpful | perpsychihelppyr | percep_psq_anual_dic | Binary recode implemented from the psychiatry helpfulness field: source categories 0–1 = 0 and category 2 = 1. Because the available labels displayed four response levels while the do-file used codes 0–2, and because the source alias is inconsistent, confirm the coding before reuse. | Exploratory outcome among 12-month psychiatry users in Supplementary Table S2B; retained with an explicit confirmation flag. |
| Psychology care perceived as very helpful | perpsychohelppyr | percep_psc_anual_dic | Binary recode of percep_psc_anual: source categories 0–1 = 0 and category 2 = 1. The available labels displayed four response levels while the do-file used codes 0–2; confirm the coding before reuse. | Exploratory outcome among 12-month psychology users in Supplementary Table S2B; retained with an explicit confirmation flag. |
| **H. Exploratory clinical-complexity indicators** |  |  |  |  |
| Recent diagnostic activity | patpsq_1 and pmtpsq_1 | actividad_30d | Defined among participants with psiq_anual = 1. Coded 0 when psiq_mensual = 0 (12-month disorder without a 30-day disorder) and 1 when psiq_mensual = 1 (12-month and 30-day disorder). It does not measure persistence, chronicity, episode duration, or severity. | Exposure and descriptive indicator in Supplementary Table S3. |
| Number of observed 12-month disorder domains | patansiedad, pacta1, prev_anual_psicosis, prev_trast_alimentacion_anual, and patundepC | n_dom_obs_anual | Row count of nonmissing values across five supplied 12-month domains: anxiety, affective, psychosis screen, eating disorder, and harmful addictive-substance use or dependence. | Quality-control input for the partial comorbidity measure in Supplementary Table S3. |
| Number of positive 12-month disorder domains | patansiedad, pacta1, prev_anual_psicosis, prev_trast_alimentacion_anual, and patundepC | n_dom_pos_anual | Row sum across the same five binary domains; set to missing unless all five domain indicators were observed. | Intermediate input for the partial comorbidity measure in Supplementary Table S3. |
| Partial five-domain comorbidity | n_dom_obs_anual, n_dom_pos_anual, and patpsq_1 | comorb_5dom_bin | Defined only among participants with psiq_anual = 1 and complete information on all five domains. Coded 0 for exactly one positive domain and 1 for two or more positive domains. This is not a complete diagnostic count. | Exposure and descriptive indicator in Supplementary Table S3. |
| **I. Legacy variables retained only for the audit trail** |  |  |  |  |
| Legacy direct 12-month need identified by others | demannose | need_others_anual | Direct copy of others_need_anual from an earlier analysis. It does not incorporate SR111 or SR113. | Retained for traceability; not used in the final SR113-based Tables 4–5. |
| Legacy direct 12-month self-perceived need | demanse | need_self_anual | Direct copy of self_need_anual from an earlier analysis. It does not incorporate contact-based routing through SR111. | Retained for traceability; not used in the final SR113-based Tables 4–5. |
| Legacy any 12-month perceived need | demanse and demannose | need_any_anual | Earlier binary summary based only on the two direct items: 1 if either direct item equaled 1; 0 if both equaled 0; otherwise missing. | Retained for traceability; not used in the final SR113-based Tables 4–5. |
| Legacy contact gap among any direct perceived need | need_any_anual and mhs_anual | bch_need_any_anual | Among participants with need_any_anual = 1, coded as 1 − mhs_anual. | Retained for traceability; not based on the final SR111/SR113-derived perceived-need measure. |
| Legacy contact gap among direct self-perceived need | self_need_anual and mhs_anual | bch_need_self_anual | Among participants with self_need_anual = 1, coded as 1 − mhs_anual. | Retained for traceability; not based on the final derived perceived-need measure. |
| Legacy contact gap among direct need identified by others | others_need_anual and mhs_anual | bch_need_others_anual | Among participants with others_need_anual = 1, coded as 1 − mhs_anual. | Retained for traceability; not based on the final SR111/SR113-derived perceived-need measure. |
| Legacy perceived-need type | demanse and demannose | need_type | Available script defined 0 = neither direct item positive, 1 = self-perceived only, and 2 = identified by others only. A downstream recode also anticipated value 3, but the assignment of value 3 was not present in the reviewed definition. | Retained only for auditability; not used in final SR113-based results and requires confirmation before reuse. |
| Legacy any perceived need | need_type | need_any | Binary recode of need_type: 0 = no perceived need and values 1–3 = any perceived need. | Retained only for auditability; not used in final SR113-based results. |

Notes. ENSM, National Mental Health Survey; CAPI, computer-assisted personal interviewing. A contact gap denotes absence of measured contact among an eligible disorder-defined subpopulation; it should not be interpreted as absence of adequate, continuous, disorder-specific, or effective treatment. Exact source-field names and Stata analysis-variable names are shown to support reproducibility. Items requiring confirmation are identified explicitly rather than resolved by inference.
